# Sequential intravenous and intracerebroventricular GD2-CAR T-cell therapy for H3K27M-mutated diffuse midline gliomas

**DOI:** 10.1101/2024.06.25.24309146

**Authors:** Michelle Monje, Jasia Mahdi, Robbie Majzner, Kristen Yeom, Liora M. Schultz, Rebecca M. Richards, Valentin Barsan, Kun-Wei Song, Jen Kamens, Christina Baggott, Michael Kunicki, Alexandria Sung Lim, Agnes Reschke, Sharon Mavroukakis, Emily Egeler, Jennifer Moon, Shabnum Patel, Harshini Chinnasamy, Courtney Erickson, Ashley Jacobs, Allison K. Duh, Skyler P. Rietberg, Ramya Tunuguntla, Dorota Danuta Klysz, Carley Fowler, Sean Green, Barbara Beebe, Casey Carr, Michelle Fujimoto, Annie Kathleen Brown, Ann-Louise G. Petersen, Catherine McIntyre, Aman Siddiqui, Nadia Lepori-Bui, Katlin Villar, Kymhuynh Pham, Rachel Bove, Eric Musa, Warren Reynolds, Adam Kuo, Snehit Prabhu, Lindsey Rasmussen, Timothy T. Cornell, Sonia Partap, Paul G. Fisher, Cynthia J. Campen, Gerald Grant, Laura Prolo, Xiaobu Ye, Bita Sahaf, Kara L. Davis, Steven A. Feldman, Sneha Ramakrishna, Crystal Mackall

## Abstract

H3K27M-mutant diffuse midline gliomas (DMGs) express high levels of the GD2 disialoganglioside and chimeric antigen receptor modified T-cells targeting GD2 (GD2-CART) eradicate DMGs in preclinical models. Arm A of the Phase I trial NCT04196413 administered one IV dose of autologous GD2-CART to patients with H3K27M-mutant pontine (DIPG) or spinal (sDMG) diffuse midline glioma at two dose levels (DL1=1e6/kg; DL2=3e6/kg) following lymphodepleting (LD) chemotherapy. Patients with clinical or imaging benefit were eligible for subsequent intracerebroventricular (ICV) GD2-CART infusions (10-30e6 GD2-CART). Primary objectives were manufacturing feasibility, tolerability, and identification of a maximally tolerated dose of IV GD2-CART. Secondary objectives included preliminary assessments of benefit. Thirteen patients enrolled and 11 received IV GD2-CART on study [n=3 DL1(3 DIPG); n=8 DL2(6 DIPG/2 sDMG). GD2-CART manufacturing was successful for all patients. No dose-limiting toxicities (DLTs) occurred on DL1, but three patients experienced DLT on DL2 due to grade 4 cytokine release syndrome (CRS). Nine patients received ICV infusions, which were not associated with DLTs. All patients exhibited tumor inflammation-associated neurotoxicity (TIAN). Four patients demonstrated major volumetric tumor reductions (52%, 54%, 91% and 100%). One patient exhibited a complete response ongoing for >30 months since enrollment. Eight patients demonstrated neurological benefit based upon a protocol-directed Clinical Improvement Score. Sequential IV followed by ICV GD2-CART induced tumor regressions and neurological improvements in patients with DIPG and sDMG. DL1 was established as the maximally tolerated IV GD2-CART dose. Neurotoxicity was safely managed with intensive monitoring and close adherence to a management algorithm.

## INTRODUCTION

Chimeric antigen receptors (CARs) couple an antigen binding domain to T-cell signaling domains to redirect T lymphocytes to cancer cells expressing a target of interest. Autologous CAR T-cells have mediated impressive results in refractory B cell malignancies and plasma cell malignancies ^1-5^, but have not demonstrated high rates of sustained antitumor effects in solid cancers or brain tumors ^6-13^ with the exception of promising responses in a recent trial of GD2-CAR T cell therapy for neuroblastoma^14^. Differential rates of activity between liquid cancers and solid/brain tumors may relate to a dearth of safe targets with high homogenous expression, inadequate T-cell trafficking, and/or T-cell dysfunction induced by the tumor microenvironment (TME).

H3K27M-mutated diffuse midline gliomas (DMGs) chiefly occur in children and young adults and originate in midline structures of the nervous system. Patients with pontine DMG (also called diffuse intrinsic pontine glioma, DIPG) have a median overall survival of 11 months from diagnosis, and 5-year overall survival <1%^15-17^. Patients with DMGs outside of the brainstem, including the spinal cord, have a median overall survival of 13 months^16^. DIPG is the most common cause of death due to brain cancer in children. Palliative radiotherapy is the standard of care. Cytotoxic chemotherapy has not improved outcomes to date^18^. While targeted therapies and immuno-oncology strategies have begun to show early promise^19-21^, outcomes remain dismal.

We discovered high, uniform expression of GD2, a disialoganglioside, on H3K27M+ DMG cells and demonstrated that IV administration of GD2-CART eradicated established DMGs in patient-derived orthotopic xenograft mouse models^22^. We and others also demonstrated increased potency and decreased systemic inflammation following intracerebroventricular (ICV) administration of CAR T-cells compared to intravenous (IV) administration in preclinical brain tumor models^23-26^. These data provided rationale for this first-in-human/first-in-child Phase 1 clinical trial (NCT04196413, Phase 1 Clinical Trial of Autologous GD2 Chimeric Antigen Receptor (CAR) T-cells (GD2-CAR T) for Diffuse Intrinsic Pontine Gliomas (DIPG) and Spinal Diffuse Midline Glioma (sDMG). We previously reported antitumor activity and correlative findings from the first three patients treated on NCT04196413 Arm A at dose level 1 (DL1) and a fourth who enrolled on trial but received therapy via a single patient compassionate IND^27^. Here we report final clinical results following enrollment completion of Arm A, which demonstrate tolerability of 1e6 GD2-CART cells/kg IV (DL1) followed by sequential ICV infusions, tumor regressions and, in some cases, sustained antitumor effects in patients with DIPG and sDMG.

## METHODS

### Trial Design

Eligible patients were 2-30 years of age with biopsy-confirmed H3K27M-mutated DIPG or sDMG, who had completed standard frontline radiotherapy at least 4 weeks prior to enrollment, were not receiving corticosteroid therapy and had acceptable performance status (≥ 60 Lansky or Karnofsky scores). While patients typically exhibited bulky disease in the brainstem or spinal cord, those with bulky disease involving the thalamus or cerebellum were not eligible due to increased risk of toxicity of GD2-CART observed with these tumor locations in murine models^22^. Patients with clinically significant dysphagia (as an indicator of significant medullary dysfunction) were also ineligible. Detailed eligibility and exclusion criteria are contained in **Supplemental Methods 1**. The clinical trial protocol was approved by the Stanford Institutional Review Board (IRB) and registered with ClinicalTrials.gov (NCT04196413). Informed patient or parent consent and child assent were obtained.

Primary objectives were to determine feasibility of manufacturing, assess safety and tolerability and identify a maximally tolerated and/or recommended phase 2 dose of IV GD2-CART following lymphodepleting chemotherapy in this population. Secondary objectives (detailed in **Supplemental Methods 2, Section 1)** included preliminary assessment of benefit as measured by radiographic response on MRI and by clinical improvement. IV doses were escalated using a 3+3 design, based upon the occurrence of dose limiting toxicities possibly, probably, or likely attributed to IV GD2-CAR T and occurring within 28 days following infusion. DLTs comprised any grade 5 toxicity, grade 4 cytokine release syndrome (CRS), grade 4 neurotoxicity lasting >96h, new grade 3 neurotoxicity lasting > 28d, grade 4 neutropenia or thrombocytopenia lasting >28d, and grade 3 non-hematologic toxicity with the exceptions of those detailed in (**Supplemental Methods 2, Section 5.4.5**). Because sDMGs are rare, the protocol allowed safety in the DIPG cohort to inform dose escalation for patients with sDMG but given the risk for neurotoxicity related to the location of DIPG, safety in sDMG patients did not inform dose escalation for DIPG patients.

In December 2020, a protocol amendment added a new secondary objective to evaluate safety and assess clinical benefit of patients treated with IV GD2-CAR T-cells followed by ICV administrations of GD2-CAR T-cells (**Supplemental Methods 2**). Patients were eligible to receive second or subsequent IV or ICV infusions if they showed CR, PR, MR, or SD radiographically on MRI, or had evidence for clinical benefit from pre-infusion baseline according to the protocol-specified clinical neurological exam, and at least 28 days had elapsed since the initial CAR T infusion or 21 days from subsequent infusions, circulating CAR T-cell levels were < 5% and toxicity had resolved to < grade 2. Lymphodepletion was not administered prior to ICV infusions. All subsequent infusions were delivered ICV.

### GD2-CART Manufacturing and Product Assessment

GD2-CART were manufactured in the automated CliniMACS Prodigy using a 7-day process incorporating IL-7 (Miltenyi Biotec, 12.5 ng/mL) and IL-15 (Miltenyi Biotec, 12.5 ng/mL) as illustrated in (**Fig. S1a)** and detailed in **Supplemental Methods 3**. GD2-CART was encoded by a retroviral vector encoding an iCasp9 domain (Bellicum Pharmaceuticals, Inc.) and GD2.4-1BB.CD3z CAR separated by a P2A ribosomal skip sequence (**Fig. S1b)**.

### Supportive Care, Toxicity Monitoring and Management

Patients received standard supportive care for seizure prophylaxis, immunosuppression associated with lymphodepleting chemotherapy, CRS and immune effector cell acute neurotoxicity syndrome (ICANS) as detailed in **Supplemental Methods 4**. Neurotoxicity that was distinct from ICANS and attributable to local tumor inflammation was designated tumor inflammation-associated neurotoxicity (TIAN)^23,28^ and graded using NCI CTCAEv5.0. To mitigate risks associated with TIAN, we implemented a toxicity monitoring and management algorithm that i) placed Ommaya catheters or similar devices for intracranial pressure monitoring and potential treatment of hydrocephalus in all patients ii) incorporated sequential neurological exams and scheduled and symptom-prompted intracerebral pressure monitoring, iii) prescribed measures to lower elevated ICP (positioning, hypertonic saline (3%), CSF removal) in patients with documented elevated ICP and iv) administered anakinra and corticosteroids in patients with significant neurologic toxicity (detailed in **Supplemental Methods 4**).

### Response Assessment and Correlative Studies

Clinical response was assessed using the Clinical Improvement Scores (CIS)^27^ generated via protocol-directed neurological examinations performed by a neuro-oncologist at prescribed times following GD2-CAR T-cell administration. The CIS represents a simple quantification of the neurological exam, which added or subtracted one point for each symptom/sign that improved or worsened respectively from the patient’s pre-infusion baseline exam (**Supplementary Methods 5**). For patients who received corticosteroid therapy for toxicity management, CIS scoring was deferred until at least 7 days after corticosteroid discontinuation. Radiographic responses were assessed with MRI scans of the brain and/or spinal cord with and without gadolinium. Because DMGs are diffusely infiltrative of CNS structures and difficult to measure in linear dimensions, volumetric segmentation of tumor corresponding to abnormal T2 signal was performed by a neuroradiologist to measure radiographic change in tumor volume, consistent with RANO 2.0 recommendations^29^. Peripheral blood and CSF samples were collected prior to and following IV and ICV infusions to measure cytokine/chemokine levels, GD2-CART persistence, and cell-free tumor DNA (ctDNA), as previously described^27^.

### Statistical Analysis

Sample size estimation was based on clinical considerations and the Phase I 3+3 design. The targeted DLT rate was at ≤30%. All patients who enrolled into Arm A and received one infusion of GD2-CAR T-cells were included in the analysis. Descriptive statistics were used to summarize baseline patient and disease characteristics, toxicity data, correlative and clinical outcomes. Tumor response rate was estimated using the binomial distribution. Overall survival was measured from date of diagnosis to date that event occurred or censored at time of data cut-off. Survival probability was estimated using the Kaplan-Meier method^30^. The confidence interval of median survival time was constructed by the method of Brookmeyer-Crowley ^31^. All comparisons made in correlative outcomes were exploratory and no adjustments were made for multiple comparisons. Statistical analyses were conducted using Prism software.

## Results

### Demographics

Enrollment began in June 2020 and the data cutoff was December 1, 2023. A consort diagram is shown in **Fig. S2**. Characteristics of the thirteen enrolled patients are shown in **Table 1**. Median age was 15 yrs (range 4-30 yrs), and seven patients were female. Ten patients had DIPG and three had sDMG. The H3K27M mutation was demonstrated in biopsies from all tumors by immunohistochemistry or DNA sequencing. Median time from diagnosis to enrollment was 5.0 mos (range 3.9-11.6). Eight patients had disease progression or pseudoprogression on MRI imaging at enrollment, while five did not have documented progression at enrollment. Two patients (Patients 002 and 011) were removed from study prior to treatment due to rapid tumor progression and a decline in performance status rendering them ineligible to protocol-directed therapy.

**Table 1.** Patient Demographics and GD2-CAR T-Cell Infusions Received.

| UPN | Age bracket/<br>Sex | Disease | Histone Mutation | Other Known Mutations and Molecular Aberrations | At Enrollment |  |  | IV | ICV |  |
| --- | --- | --- | --- | --- | --- | --- | --- | --- | --- | --- |
|  |  |  |  |  | Mos since Dx | Mos since end of XRT | Post-XRT Progression | Dose (x10<sup>6</sup>/kg) | Dose (x10<sup>6</sup>) | # doses |
| 001 | 11-15/F | DIPG | H3.3H27M | TP53, KIT amplification, MET amplification, and NBN rearrangement | 9.9 | 8 | Yes | 1 | n/a | 0 |
| 003* | 21-25/M | DIPG | H3.3H27M | TP53, ATRX, RB1 | 15.5 | 12.5 | Yes | 1 | 30 | 4 |
| 004* | 0-5/F | DIPG | H3.3H27M | TP53, DAXX rearrangement | 13.3 | 11.5 | No | 1 | 30 | 2 |
| 005 | 11-15/F | DIPG | H3.3H27M | NF1 and PIK3CA | 4.5 | 1.6 | No | 2 | 30 | 4 |
| 006 | 26-30/F | DMG | H3K27M & | From cell-free tumor DNA in CSF: 1. TP53, BRCA1, NBN, NF1, MGA rearrangement, CASP8 rearrangement, RAF1 rearrangement: | 5.1 | 1.5 | Yes | 2 | 30 | 8 |
| 007 | 0-5/F | DIPG | H3.3K27M | KDM5A, PTEN | 4.1 | 2 | No | 2 | 10, 30, 50 | 16 |
| 008 | 16-20/M | DIPG | H3K27M & | TP53 | 4.6 | 2 | Yes | 2 | 30 | 1 |
| 009 | 26-30/M | DMG | H3K27M & | patchy ATRX expression | 5.7 | 1.6 | Yes | 2 | 10, 30 | 11 |
| 010 | 16-20/M | DIPG | H3.3K27M | NF1 | 4.1 | 1.5 | Yes | 2 | 30, 50 | 15 |
| 012 | 11-15/F | DIPG | H3.3K27M | TP53, FANCM, MYD88, TSC2 | 3.9 | 1 | No | 2 | 30 | 1 |
| 013 | 6-10/M | DIPG | H3.3K27M | TP53 | 4.9 | 2 | Yes | 2 |  | 0 |

&determined by IHC; Patients 003 and 004 were previously reported as “DIPG Patient 2” and “DIPG Patient 3”, respectively, in Majzner et al., 2022<sup>27</sup>. Note that Patients 002 and 011 became ineligible after enrollment and before infusion; Patient 002 was treated on an eIND and was reported as “spinal cord DMG patient #1” in Majzner et al., 2022<sup>27</sup>.

### Manufacturing feasibility and characterization of cell products

All enrolled patients had GD2-CART products successfully manufactured that met IV or ICV dose levels. Mean manufacturing duration was 7 days. The median time from enrollment to IV GD2-CART infusion was 22.9 days (range: 15-38) days. Products demonstrated a mean fold-T cell expansion of 21.97 (range 12.16 -28.06), mean percent viability of 93.51 (range 86.8-96.9), mean transduction efficiency of 57.27 (range 21.63 – 83.80) **(Supplemental Table 1)**, and mean vector copy number per cell (VCN/CAR+ cell) of 3.17 (range 0.84 – 7.2)**(Fig. S3C,D)**. Detailed GD2-CART phenotypic composition revealed a predominance of central memory T cells (**Fig. S1C-F)** and functional assessment demonstrated GD2-specific reactivity (**Fig. S3A,B).**

### Treatment Received and Toxicity

Of eleven patients who received one IV GD2-CART infusion, nine experienced clinical or imaging benefit on MRI and received additional ICV infusions as noted in **Table 1** (n=4 median, range 1-16 infusions) administered over 1 to 29.4 months. Incidence and grade of CRS, ICANS and TIAN following each IV and ICV infusion is shown in **Fig 1a**, **Table 2, Table S2 and S3**. Following IV GD2-CART, all patients experienced CRS, with 1 of 3 patients on DL1 experiencing grade 2 CRS, 6 of 8 patients on DL2 experiencing ≥ grade 2 CRS and 3 patients experiencing DLT attributed to grade 4 CRS (**Table 2**). After IV GD2-CART, we observed ICANS in one of three patients at DL1 (grade 2) and 4 of 8 patients at DL2 (n=1 grade 3, n=3 grade 1). Based upon these results, we identified DL2 (3 x 10e6 GD2-CART/kg) as exceeding the maximally tolerated dose for IV administration in patients with DIPG/DMG. Among 62 ICV infusions, no DLTs occurred. Forty-one ICV infusions (66%) were associated with no CRS. Among 19 ICV infusions associated with CRS, most were low grade (n=16 Grade 1, n=4 infusions in 2 patients Grade 2, n=1 Grade 3 in the context of urosepsis in a patient with sDMG) (**Table 2**). No ICANS was observed following ICV infusions.

**Figure 1.**
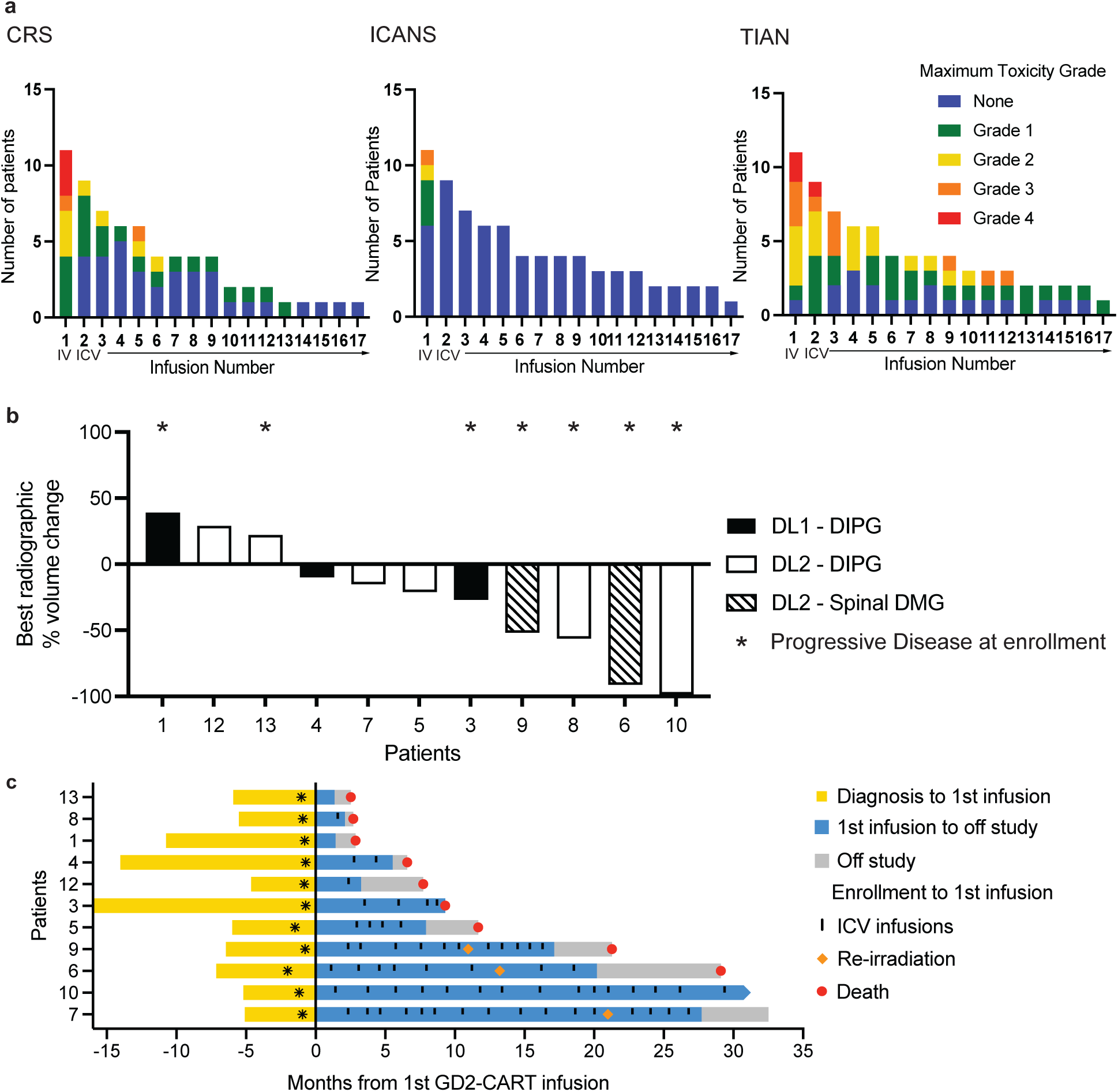
Toxicity and response measures. **a)** The number of patients experiencing CRS (left graph), ICANS (middle graph) and TIAN (right graph) following each GD2-CAR T infusion is shown. Grade of maximal toxicity indicated by color as shown in the legend. Infusion 1 was administered IV, while infusions 2-13 were administered ICV. **b)** Waterfall plot depicting maximal volumetric change in tumor volume from baseline obtained prior to first infusion. Asterisks (*) indicate those patients with documented disease progression at the time of first CAR T infusion. Black = DIPG patients treated at DL1, white = DIPG patients treated at DL2, hatched marks = spinal DMG patients treated at DL2. **c)** Swimmer’s plot depicting patient survival. Each bar represents the time from diagnosis to first treatment (yellow), time on trial (blue) until death (red dot) or data cutoff for individual patients. The time interval following removal from study for disease progression to death is depicted in grey. Vertical marks indicate each ICV infusion. Asterisks indicates time of trial enrollment, with first treatment indicated by the y-axis at time 0. Pause in infusions for focal therapy was allowed per protocol, and two patients (Patients 006 and 009) received re-irradiation late in their course, as indicated by orange diamond. Imaging and clinical benefit in Patients 003 and 004 was previously reported^27^; please note when comparing this report with the previous report^27^ that Patient 003 here = “DIPG Patient 2” in the previous report^27^ and Patient 004 here= “DIPG Patient 3” in the previous report^27^.

**Table 2.**
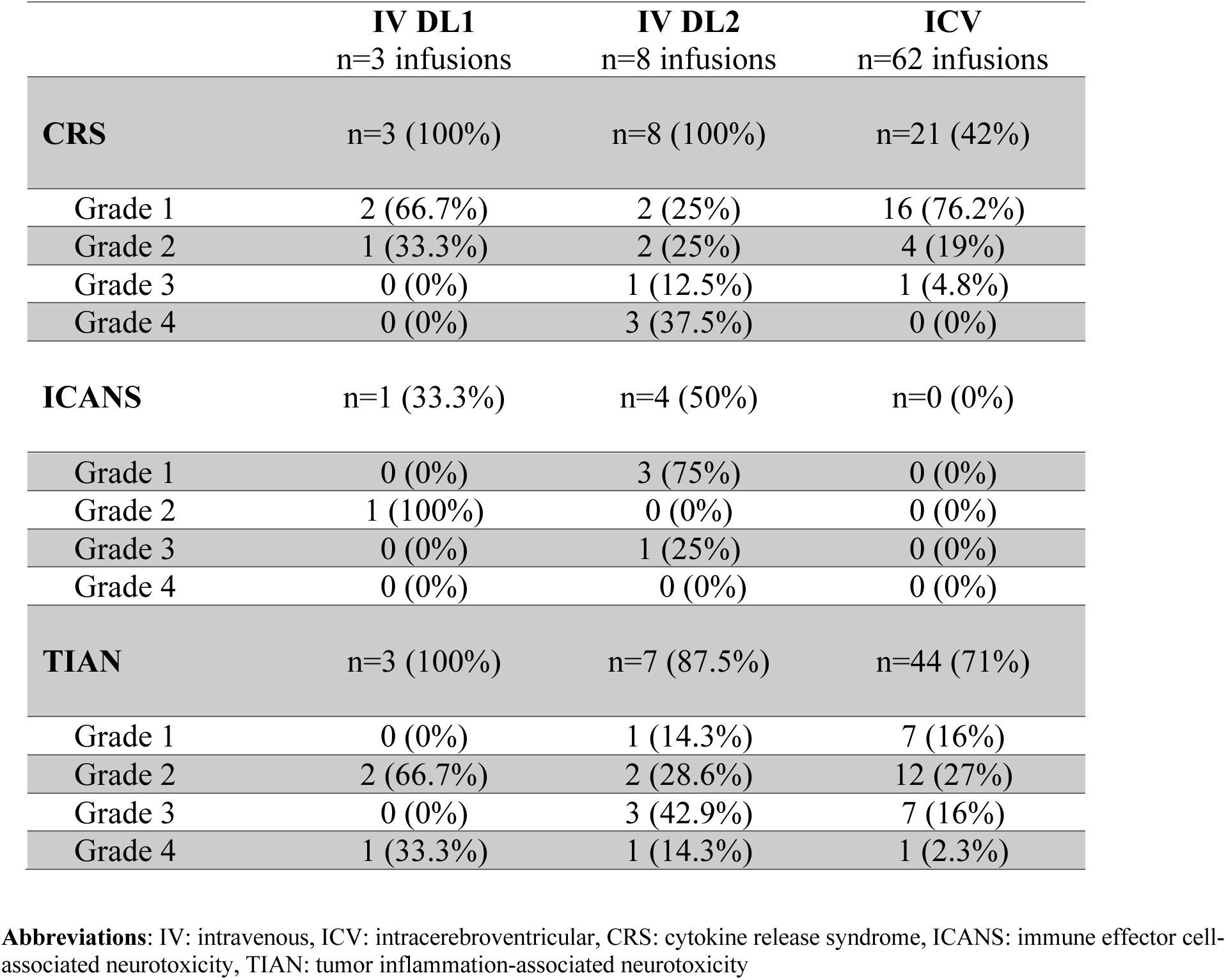
Maximum Toxicity Grades After GD2-CAR T Therapy.

We observed TIAN in 91% of patients following IV infusion and 100% of patients following the first ICV infusion. The TIAN grade typically diminished with subsequent infusions (**Fig. 1a)**. Forty-four ICV infusions (71%) were associated with TIAN. Nine of 9 (100%) patients with DIPG developed TIAN following IV GD2-CART (n=2 grade 4, n=3 grade 3, and n=4 grade 2), and 67% of patients with DIPG developed TIAN following ICV GD2-CAR T-cell infusions (n=29/43 infusions with TIAN: grade 4 n=1, grade 3 n=3, grade 2 n=9, grade 1 n=16). TIAN reversed in all patients following treatment according to our toxicity management algorithm and no patient experienced a DLT due to TIAN. In one sDMG patient, high-grade communicating hydrocephalus was observed during peak tumor inflammation. The agent available to ablate the CAR T cells via induction of caspase-9, AP1903, was not administered to any patients treated on Arm A.

### Response

Experience with the first three patients enrolled suggested a hypothesized risk of progression approximately 2-3 months following IV infusion^27^, including Patient 003 who showed an impressive response to IV infusion, followed by progression at approximately Day 70, which responded to ICV infusion. Based upon this experience, beginning with Patient 004, the protocol was amended to allow patients with clinical or imaging benefit following IV infusion to receive sequential ICV infusions. Patients were eligible to receive ICV infusions every 1-3 months as long as they had experienced stable disease or clinical or imaging benefit from enrollment.

Several patients exhibited reduction in tumor size following GD2-CART, with maximal volumetric change in tumor size for each patient shown in **Fig 1b** and overall survival from time of diagnosis to trial enrollment, and from first GD2-CART administration to time off trial, death, or the data cutoff shown in **Fig 1c**. Patient 010 with DIPG who was enrolled with evidence of progression or possibly peudoprogression by imaging following completion of upfront radiotherapy demonstrated complete response by MRI imaging sustained for >30 months following enrollment and ongoing at the time of data cutoff (**Fig 1b, 2a,b)**. Patient 006 (sDMG) demonstrated 91% reduction in tumor volume (**Fig 1b, 3a,b)**. Changes in tumor volume over time for additional patients is shown in **Fig S4a**.

**Figure 2.**
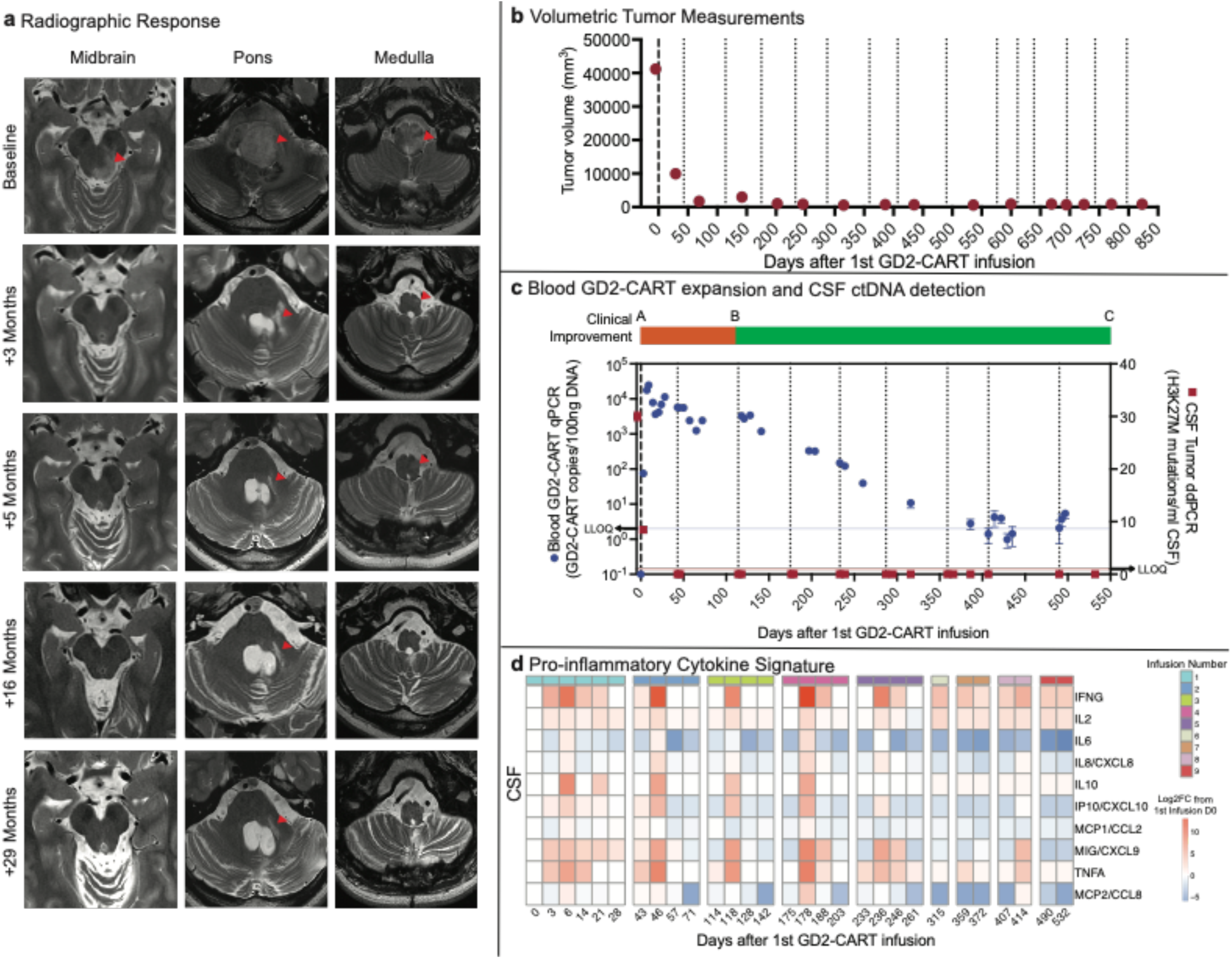
Patient 010 therapeutic response and correlative findings. **a)** T2-weighted, axial MRI images of tumor at levels of the midbrain, pons and medulla at baseline, 3 months, 5 months, 16 months and 29 months following first infusion. Red arrowheads indicate T2 signal abnormality. At baseline, extensive tumor involving the midbrain (left > right), pons and medulla is evident. Note the mass obscuring the fourth ventricle at baseline that resolves by 3 months, revealing the fourth ventricle. At 3 months, return of CSF around the brainstem is evident as the size of the brainstem normalizes, and the T2 signal normalizes throughout much of the brainstem. At 3 months, T2 signal abnormality likely representing tumor remains around the area of the biopsy tract in the pons (red arrowhead), and resolves by 5 months. At 5, 16, and 29 months, the arrowheads show stable T2 signal abnormality of left-sided pontine biopsy tract and stable area of subtle T2 hyperintensity of unclear significance in the medulla. (As is standard for axial MRI images, the patient’s left is the reader’s right). **b)** Tumor volume as a function of days following first GD2-CAR T infusion. **c)** Overlay of clinical change, CAR T-cell persistence and tumor cell-free DNA. Clinical improvement is depicted as a bar at the top of the panel, with red indicating clinical worsening from baseline and green indicating clinical improvement from baseline. This patient experienced transient worsening of symptoms followed by clinical improvement from pre-treatment baseline. CAR T-cell persistence evident in blood, as measured by qPCR of the CAR construct, is shown in blue data points (left y-axis). Cell-free tumor DNA in CSF, measured as H3K27M mutations detected by ddPCR, is shown in red data points (right y-axis). GD2-CAR T infusions indicated as dotted vertical lines. **d)** CSF cytokine levels after each infusion, expressed as log fold change from baseline prior to the first infusion.

**Figure 3.**
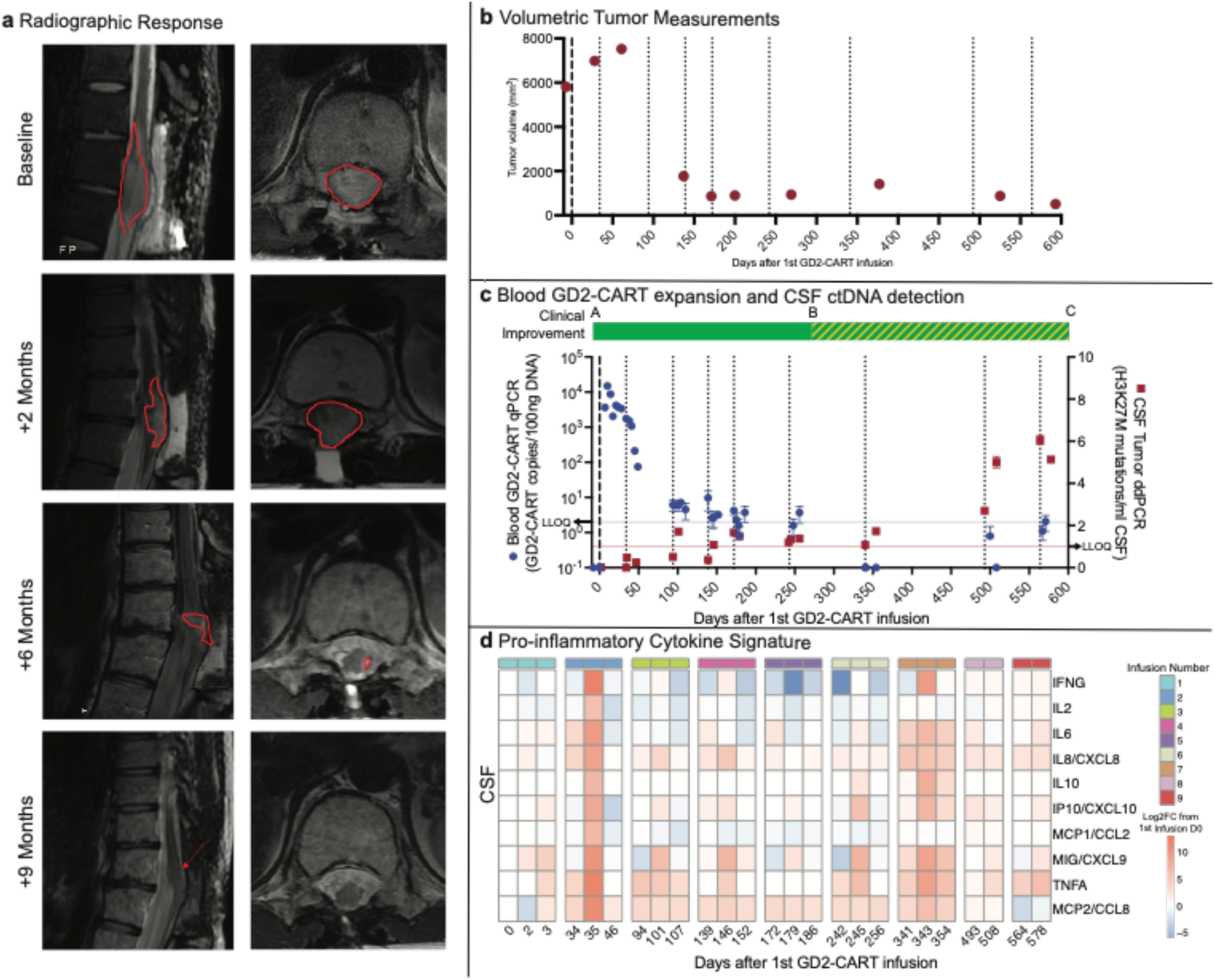
Patient 006 therapeutic response and correlative findings. **a)** T2-weighted, sagittal (left) and axial (right) MRI images of spinal cord tumor a at baseline, 2 months, 6 months and 9 months following first infusion. Red outline indicates T2 signal abnormality (tumor). At baseline, tumor centered at T11/12 spinal level diffusely involves the spinal cord and expands the cord to fill the entire spinal canal, with no CSF visualized around cord. The tumor infiltration of the spinal cord progressively improves until the cord is of normal caliber and the T2 signal abnormality is minimal (red arrow). **b)** Tumor volume as a function of days following first GD2-CAR T infusion. **c)** Overlay of clinical change, CAR T persistence and tumor cell-free DNA. Clinical improvement is depicted as a bar at the top of the panel, with green indicating clinical improvement from baseline and green hatched indicating improvement from pre-treatment baseline but worsened from peak improvement, but still improved from pre-treatment baseline. CAR T-cell persistence in blood, as measured by qPCR of the CAR construct, is shown in blue circles (left y-axis). Cell-free tumor DNA in CSF, measured as H3K27M mutations detected by ddPCR, is indicated by red squares (right y-axis). GD2-CAR T infusions indicated as dotted vertical lines. Note that as CAR T-cells become undetectable in blood by qPCR, cell-free tumor DNA elevates. This inflection point correlates with the beginning of disease progression. **d)** CSF cytokine levels after each infusion, expressed as log fold change from baseline prior to the first infusion.

Clinical improvements as measured by changes in CIS were also observed (**Supplemental Table 4, Fig S7)**. In general, clinical improvement coincided with tumor response on MRI (**Fig 2C**, **Fig 3C**). For example, prior to first infusion, patient 006 (sDMG) had severe paraplegia, neuropathic pain, and bowel and bladder dysfunction. At the time of her best overall response (>90% tumor reduction), she experienced full bowel and bladder continence, markedly improved pain, and improved lower extremity function enabling ambulation with a cane. Similarly, patient 010 (DIPG) required assistance with gait at enrollment (used a wheelchair for longer distances), but experienced a complete response by imaging accompanied by significant clinical improvement, including improved left-sided hearing, hemifacial, hemibody and taste sensation, improved motor coordination, and improved gait to the point of independent ambulation. For two patients, however, the relationship between radiographic findings on MRI and clinical improvement was not direct, with patient 007 demonstrating sustained clinical improvement without significant radiographic MRI changes and patient 005 demonstrating significant improvement in tumor size on MRI without obvious clinical benefit (**Fig. S4a, Fig. S7,** and **Table S4**). Median OS for patients treated on Arm A was 20.6 months from diagnosis, with 2 patients with DIPG alive and followed after data cutoff (patient 007: length of follow-up 33.5 mos and patient 010: length of follow-up 30.9 mos). For patients with DIPG, the median OS was 17.6 months and for sDMG the median OS was 31.96 months (**Fig. S4B)**; comparison to historical controls is not possible in this study due to the highly selected nature of the trial cohort.

### Correlative findings

Peripheral blood demonstrated GD2-CART expansion following IV infusion to levels similar to that seen in other active CAR T trials^32-34^, which persisted during ICV infusions (**Figs 2C, 3C, S5A)** but decreased over time. CSF also exhibited GD2-CART expansion after multiple repeated infusions (**S5B)** Patient 006 (**Fig 3C**) and Patient 009 (**Fig. S4a,S5A)** experienced loss of detectable GD2-CART in the peripheral blood, as measured by PCR, temporally correlated with clinical and/or imaging progression (**Table S4**). Increased cytokine/chemokine levels were present in peripheral blood following IV GD2-CART, whereas cytokine/chemokine levels were more pronounced in CSF following ICV GD2-CART, and we observed that sequential ICV CAR T-cell infusions induced sequential, transient elevations in CSF cytokine/chemokine levels (**Fig. 2D, 3D, S6**). Cell-free tumor DNA (ctDNA) in CSF as measured by digital droplet PCR (ddPCR) demonstrated decreased H3K27M mutations/mL CSF in some patients associated with tumor regression, and increased levels at times of peak inflammation or when patients experienced clinical and/or imaging progression (**Fig 2C, 3C, S5A)**. Taken together, these data demonstrate evidence for sustained expansion and persistence of GD2-CART in the blood of patients with DIPG and sDMG associated with clinical and biological evidence of antitumor activity and begin to identify potential contributors to or correlates of loss of durable response in patients.

## Discussion

Recent advances in fundamental understanding of DMG have begun to translate to clinical advances^19-21,35-37^. The present study establishes GD2-CAR T-cell therapy as a promising modality for a historically lethal CNS cancer. The rate of clinical improvements and tumor regressions on MRI imaging, including a complete response by RANO 2.0 criteria sustained for over 30 months and through the time of this report, is reason for cautious optimism, both for DIPG/sDMG therapy and more broadly for CAR T-cell therapy of solid tumors. Therapeutic activity was clearly demonstrated in several patients based upon tumor regression by imaging and improvements in neurological symptoms. Whether GD2-CAR T cell therapy favorably impacts overall survival is difficult to discern given that the eligibility criteria excluded patients with lower performance status, the need for steroid therapy, bulky thalamic or cerebellar tumor involvement, and other factors. In addition, multiple patients demonstrated MAPK pathway and ATRX/DAXX mutations, which can be associated with relatively longer survival^38^ and more localized disease^39^. Effects of GD2-CAR T cell therapy on overall survival will need to be assessed in a future phase II trials.

Cytokine release syndrome was dose-limiting following IV infusion of GD2-CART for DIPG/DMG. In contrast, CRS did not commonly complicate ICV infusions. Unexpectedly, ICANS was relatively rarely seen, and observed only after IV administration, but not with ICV infusions despite the higher levels of cytokines/chemokines evident in CSF after ICV GD2-CART infusions. Inflammation of neural structures can cause edema with consequent mechanical complications such as obstruction of CSF flow, and immune signaling can also affect neuronal function. Concordantly, varying grades of TIAN^28^ occurred commonly, as would be expected with inflammation of tumors located in structurally constrained and functionally eloquent neuroanatomical areas such as the brainstem and spinal cord. TIAN was transient and manageable with intensive support. The observed decrease in TIAN severity with repeated infusions is notable and could be due to decreased tumor burden with repeated infusions, increased immune-modulatory or immune-suppressive mechanisms at play in the tumor microenvironment, or both.

Given the promising results noted on Arm A but with dose-limiting CRS following IV administration at high dose levels, we have now initiated Arms B and C, to test ICV-only administration with and without lymphodepleting chemotherapy. Given the burden of monthly infusions on the patient, we have also included formalized assessment of quality of life and patient-reported outcomes to Arms B and C. Ongoing and future work will define the best route of administration, the role of lymphodepleting chemotherapy, and possible combination strategies to optimize this promising therapy to achieve more complete and durable responses for children and adults with H3K27M-mutated diffuse midline gliomas.

## Data Availability

All data produced in the present study are available upon reasonable request to the authors

## Acknowledgements

The authors are deeply thankful to the patients and their families who participated in this clinical trial. The authors also acknowledge Bellicum Pharmaceuticals for providing the GD2-CAR vector and AP1903. The authors thank the Stanford Human Immune Monitoring Core (HIMC) for performing cytokine assays and the Cellular Therapy Facility at Stanford for their support of this Phase I clinical trial, including cell pharmacy support. Funding for the clinical trial was provided by grants from Parker Institute for Cancer Immunotherapy, CureSearch, National Cancer Institute (R01CA263500) and the California Institute for Regenerative Medicine (CLIN2-12595). This work was also supported in part by grants from the St. Baldrick’s Foundation to the EPICC (Empowering Immunotherapies for Children’s Cancers Cancer) Team and Alex’s Lemonade Stand Foundation. We acknowledge generous funding by the Virginia and D.K. Ludwig Fund for Cancer Research (M.M. and C.L.M.), ChadTough Defeat DIPG Foundation (M.M., J.M.), and Oscar’s Kids Foundation (M.M.). R.G.M. was the Taube Distinguished Scholar for Pediatric Immunotherapy at Stanford University School of Medicine and was supported by the Be Brooks Brave Fund St. Baldrick’s Scholar Award.

## Competing interests

M.M., R.G.M., C.M., and C.L.M., are coinventors on a patent for the use of GD2 CAR T-cells for H2K27M gliomas coordinated through Stanford University. C.L.M is a coinventor on patents for the use of dasatinib and other small molecules to modulate CAR function and control CAR-associated toxicity. C.L.M. holds equity in CARGO Therapeutics, Link Cell Therapies, and GBM Newco which are developing CAR-based therapies, and consults for CARGO, Link, Immatics, Ensoma and GBM NewCo. She receives research funding from Lyell and Tune Therapeutics. R.G.M. is a cofounder of and holds equity in Link Cell Therapies and CARGO Therapeutics, and is a consultant for Lyell Immunopharma, NKarta, Arovella Pharmaceuticals, Innervate Radiopharmaceuticals, Aptorum Group, Gadeta, FATE Therapeutics (Data and Safety Monitoring Board) and Waypoint Bio. S.A.F. holds several patents in the field of cellular immunotherapy. S.P.R holds equity in Lyell Immunopharma. M.M. hold equity in MapLight Therapeutics and CARGO Therapeutics.

---

**Supplemental Figure 1.**
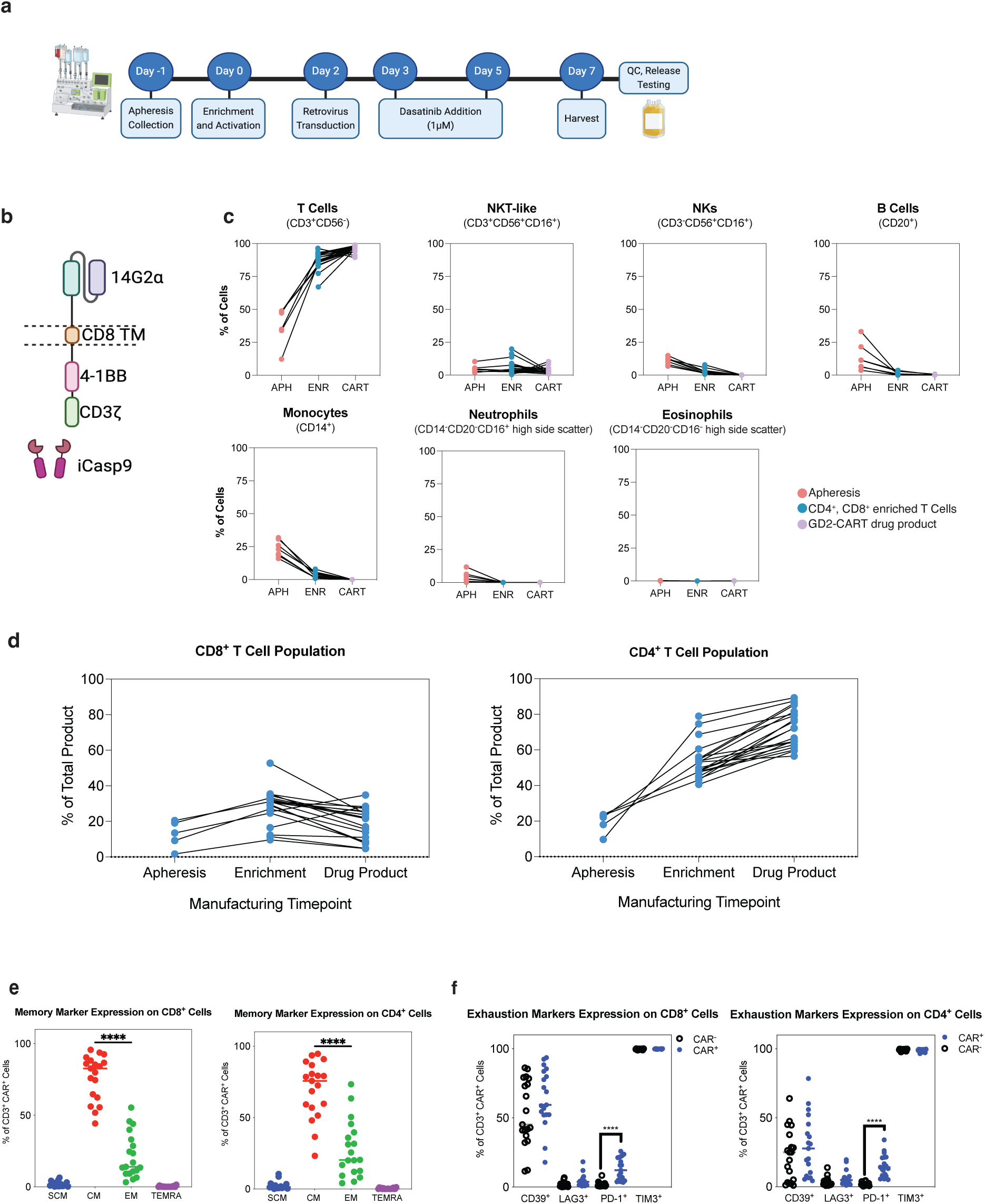
CAR T manufacturing process and drug product characterization. **a.** GD2-CART manufacturing workflow from patient apheresis collection to final drug product harvest across 7 days. **b.** Schematic representing GD2.4-1BB.CD3ζ CAR containing 14G2α binding domain, CD8α hinge and transmembrane (TM) domain, 4-1BB co-stimulatory domain, and a CD3ζ domain. **c.** Flow cytometric immunophenotyping of apheresis, CD4^+^/CD8^+^ enriched cells, and final drug product demonstrates a predominance of CD3+ T-cells and depletion of other subsets. **d.** The CD4:CD8 ratio was 1.63 ± 1.19 (SD) at apheresis, and 2.48 ± 0.06 (SD) in the GD2-CART final drug product. **e.** Phenotyping of T-cell subsets shows a predominance of central memory (CM) T-cells (CD45RA^-^CCR7^+^), followed by effector memory (EM) cells (CD45RA^-^CCR7^-^), in both CD8^+^ (left) and CD4^+^ (right) subsets in the final drug product. **f.** Exhaustion marker expression on CAR^+^ and CAR^-^ populations reveal no significant differences in CD39, LAG3, and TIM3 expression, while a significantly greater percentage (p < 0.0001, unpaired t-test) of CAR^+^ cells express of PD-1.

**Supplemental Figure 2:**
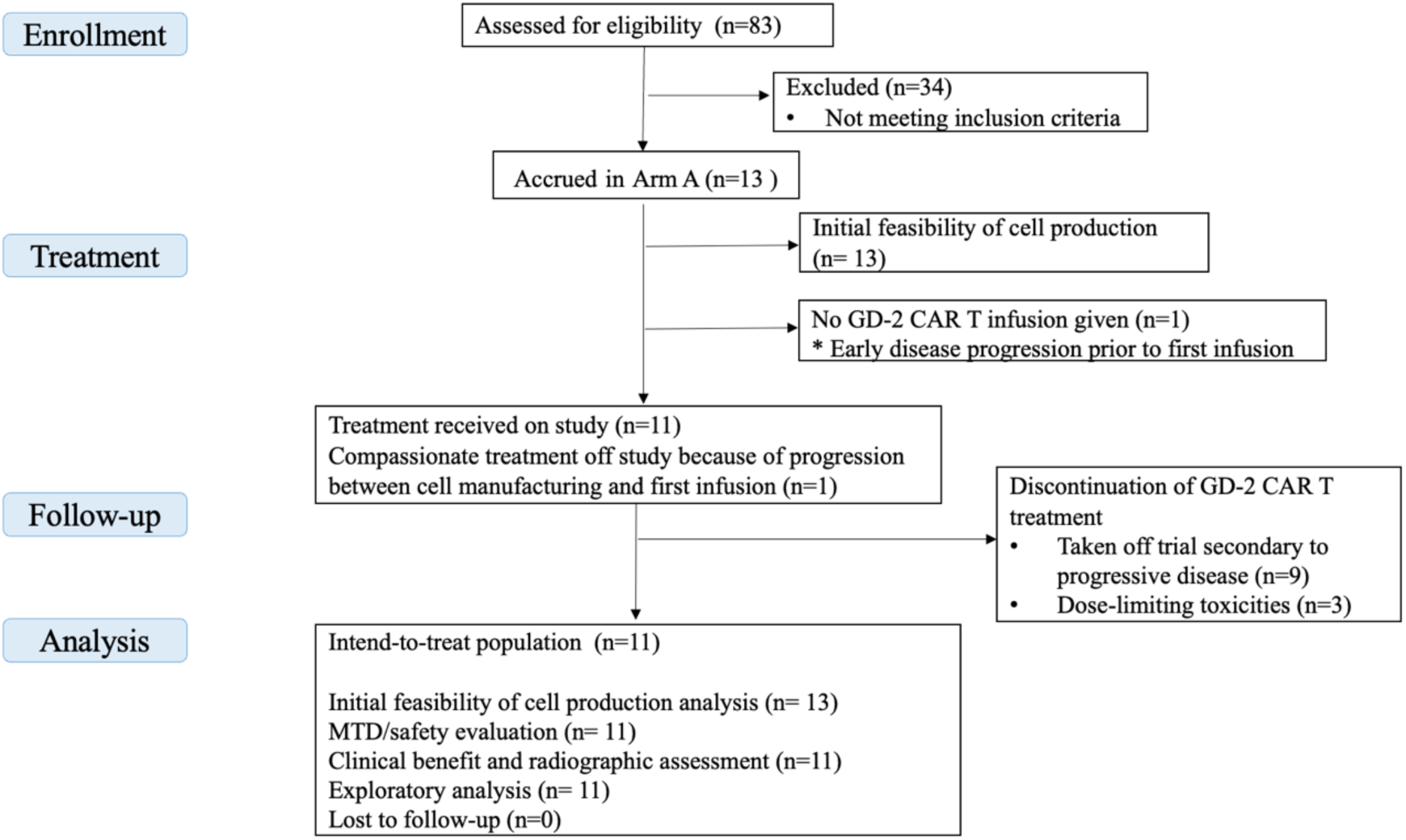
CONSORT diagram.

**Supplemental Fig. 3.**
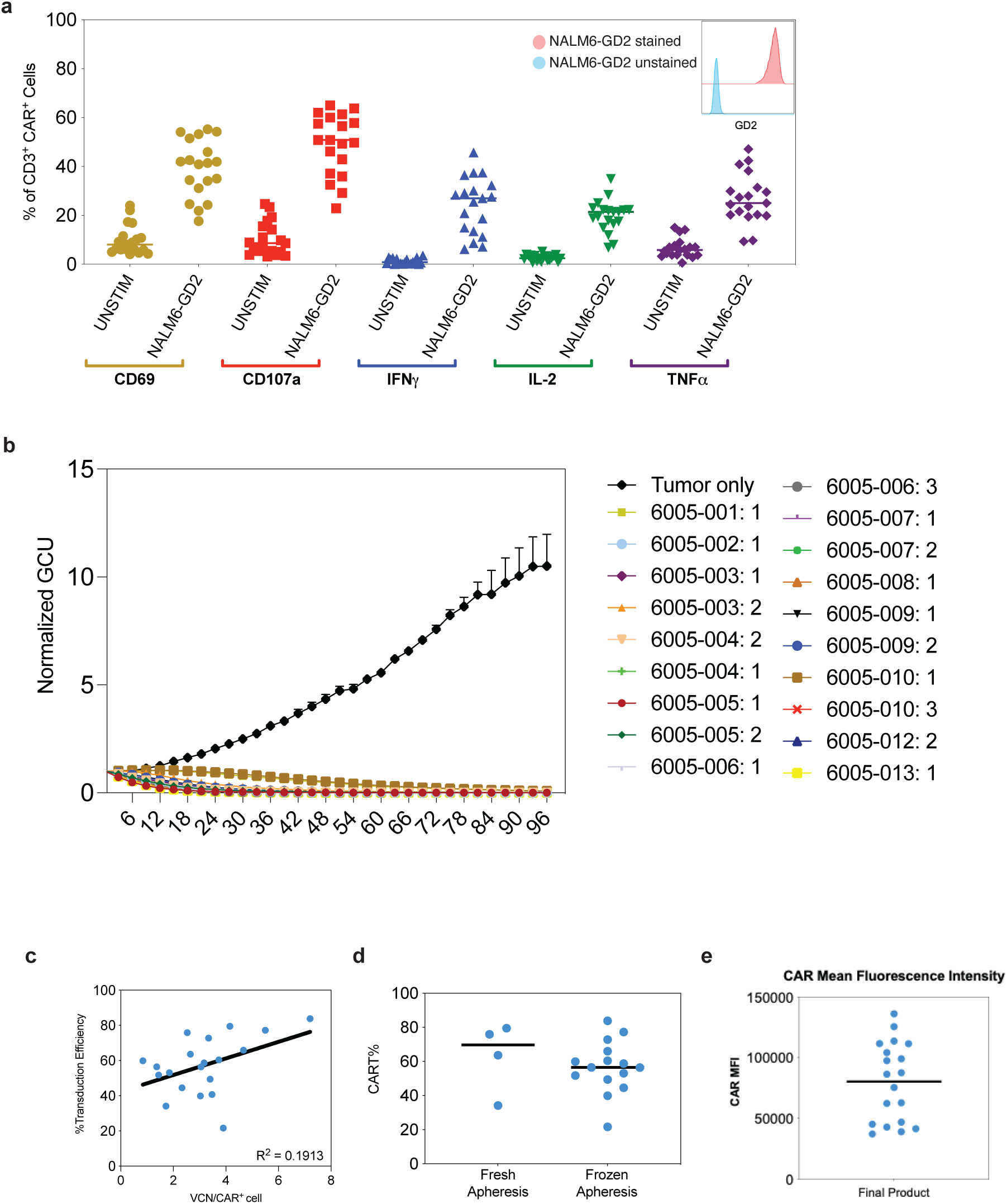
Functional assessment of GD2-CAR T-cells *in vitro*. **a**. Inset: GD2 expression on NALM6-GD2 cell line showing 38,379 and 0.5 molecules per cell, for stained (red) and unstained (blue) populations, respectively. CAR T-cell reactivity to NALM6-GD2 tumor cells measured by intracellular cytokine secretion (ICS) shows upregulation of CD69, CD107a and secretion of interferon-γ (IFNγ), interleukin-2 (IL-2) and tumor necrosis factor-α (TNFα). **b**. Incucyte based cytotoxic activity (E:T 1:1), showed that each patient drug product-controlled tumor cell growth within 96 hours of co-culture. **c.** Transduction efficiency for each patient product plotted as a function of VCN per CAR^+^ cell (R^2^ = 0.1910). **d**. No significant difference (p < 0.05, unpaired t-test) was observed in GD2-CAR transduction efficiency when manufacturing runs were executed using fresh or frozen patient apheresis material. **e.** CAR mean fluorescence intensity (MFI) values for CAR T cell expression.

**Supplemental Figure 4.**
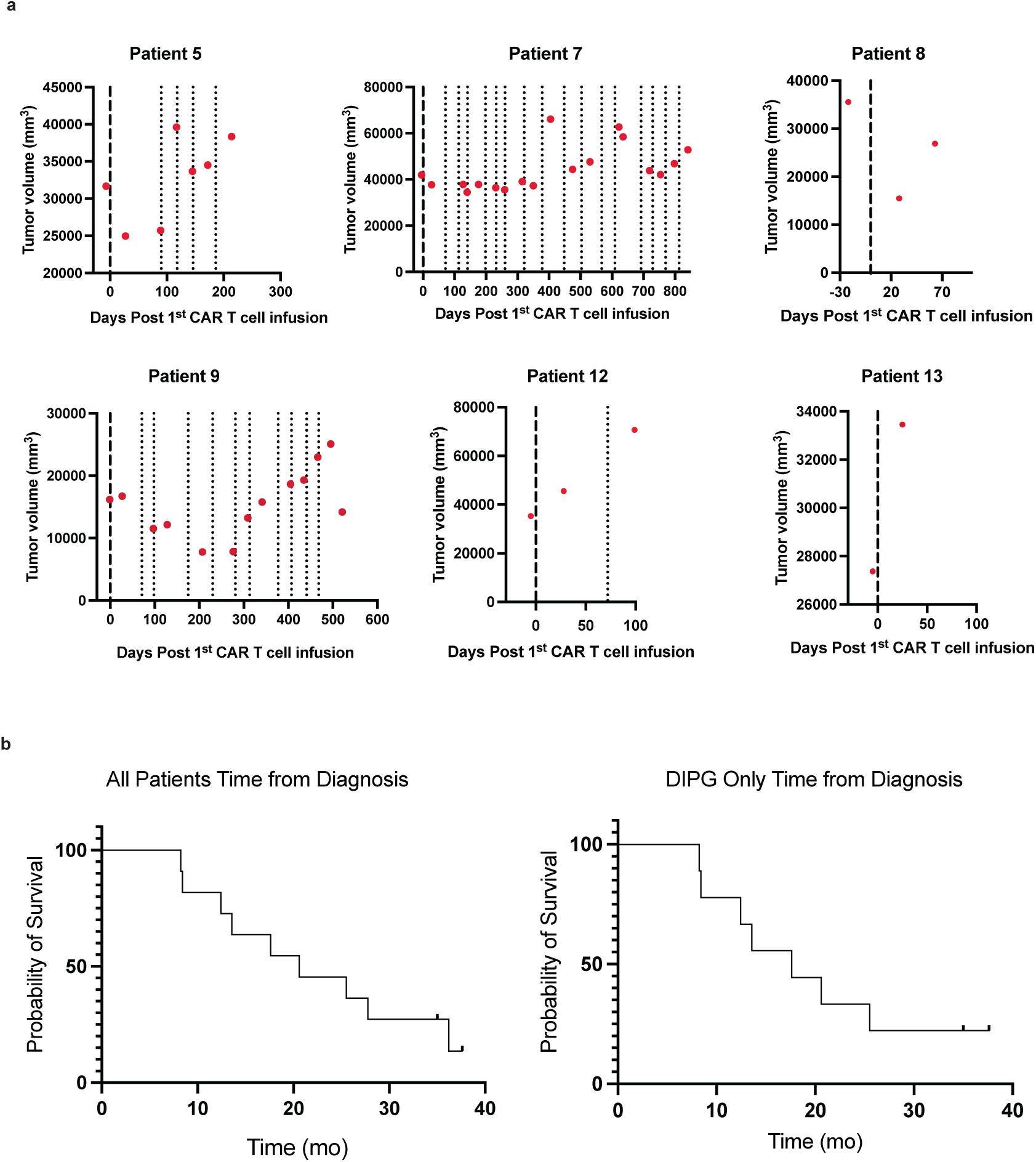
Tumor volumetric change over time and overall survival. **a.** Tumor volume as a function of days following first GD2-CART infusion is shown using the same methods used in Fig 2 and 3. Red dots represent timing of measurements; dashed line denotes IV GD2-CART infusion; dotted lines denote ICV GD2-CART infusions. Volumetric data for patients 1, 3 and 4 were presented previously in ^27^, denoted in that report as DIPG Patient #1, DIPG Patient #2 and DIPG Patient #3, respectively. Volumetric data for patients 6 and 10 are presented in Figures 2 and 3. **b.** Kaplan-Meier survival curves depicting time from diagnosis to death or data cutoff for all patients (left graph) and those patients with DIPG (right graph, excluded the two patients with spinal cord DMGs).

**Supplemental Fig. 5.**
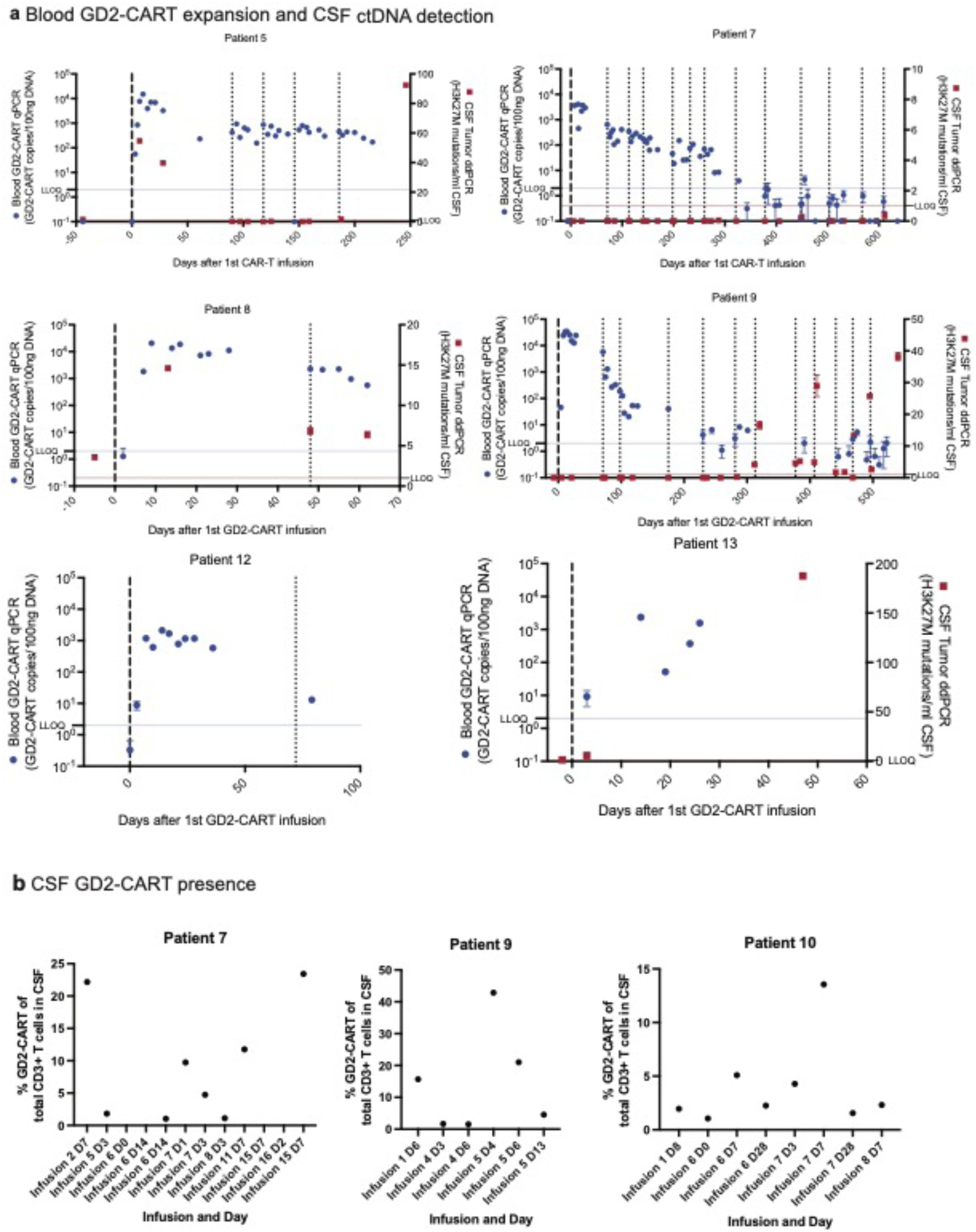
GD2 CART levels in blood and CSF, and tumor cell free DNA in CSF. **a.** GD2-CAR T copy number was detected using a validated qPCR method and based on a standard curve for CD2-CAR. The average CAR T copy number were normalized to average albumin copies by plate. Copies per 100 ng DNA were depicted as mean ± standard deviation is depicted in blue. We quantified the H3K27M mutation copies per ml of cell free CSF using the ddPCR protocol established previously ^27^ depicted in red rectangles are the average of quadruplicate mutation number/ml ± standard deviation is depicted in red. The dashed vertical lines designate infusion time points. Bold designates the initial infusion. **b.** Percent of CD3+ T cells in CSF that are GD2-CAR T cells. Flow cytometry (FACS) for GD2-CAR, using an anti-idiotype antibody, demonstrated presence of GD2-CAR T-cells in patient CSF following CAR T-cell infusions in CSF samples available for CAR-FACS assessment.

**Supplemental Fig 6:**
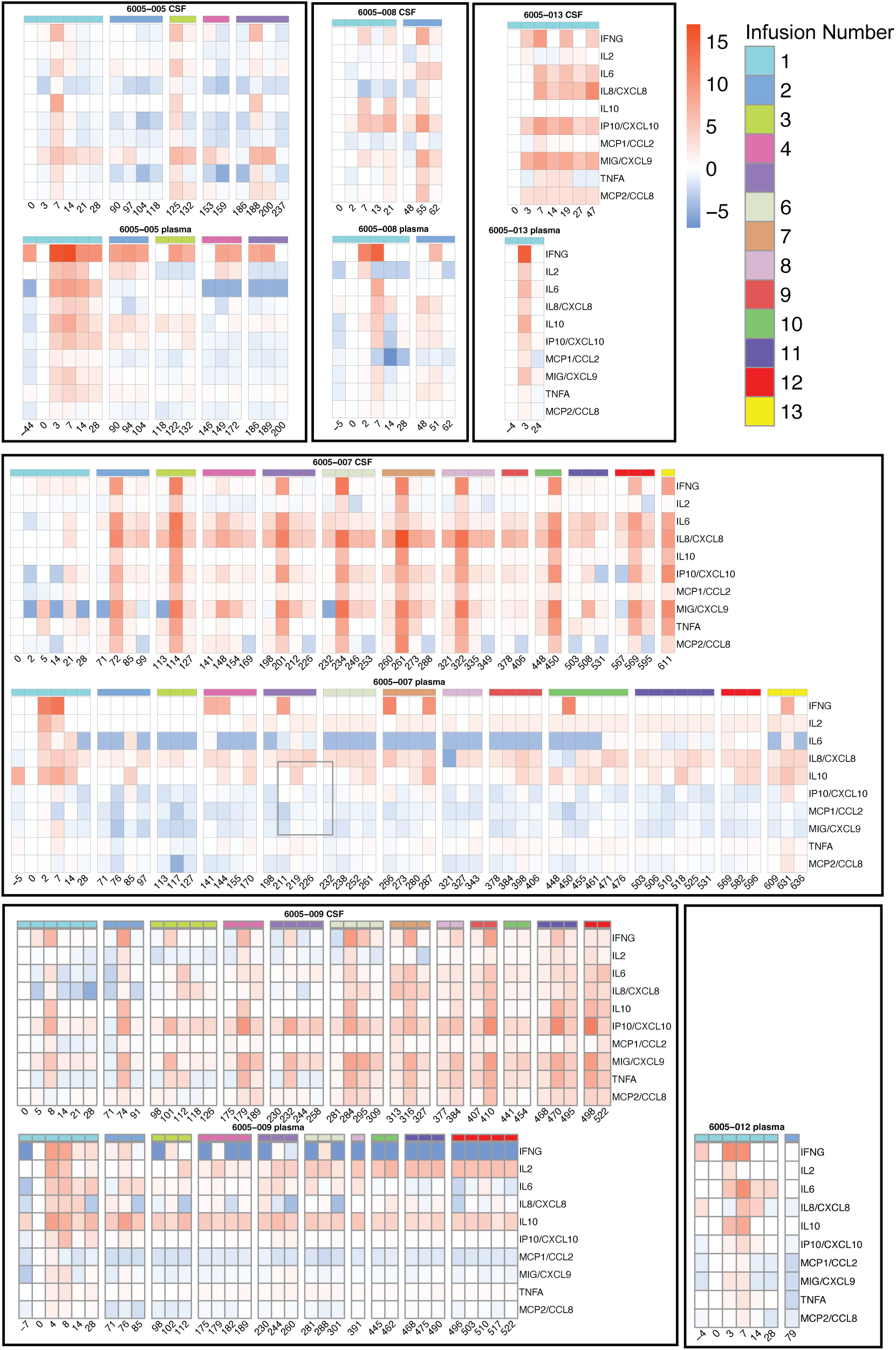
Plasma cytokine and CSF cytokines levels following GD2-CART infusions. Absolute values of inflammatory cytokines in pg/mL as a function of days following first GD2-CAR T infusion. Each individual plot represents the indicated cytokine in pg/mL as a function of days following first GD2-CAR T infusion in the indicated patient. Plots are organized by patient in columns and cytokine in rows. Each point represents the mean of technical duplicate for samples performed in technical duplicate or a single measurement. Error bars representing standard error of the mean (SEM) are displayed for samples run in technical duplicate. Dotted vertical lines represent day of an infusion. Values below the LLOQ (lower limit of quantification) are shown at the LLOQ. Values above the ULOQ (upper limit of quantification) are shown at the ULOQ and denoted by an asterisk. Each individual plot represents the absolute level of the indicated cytokine in pg/mL as a function of days following first GD2-CAR T infusion in the indicated patient. Plots are organized by patient in columns and cytokine in rows. Each point represents the mean of technical duplicate for samples performed in technical duplicate or a single measurement. Error bars representing standard error of the mean (SEM) are displayed for samples run in technical duplicate. Dotted vertical lines represent day of an infusion. Values below the LLOQ (lower limit of quantification) are shown at the LLOQ. Values above the ULOQ (upper limit of quantification) are shown at the ULOQ and denoted by an asterisk.

**Supplemental Fig 7:**
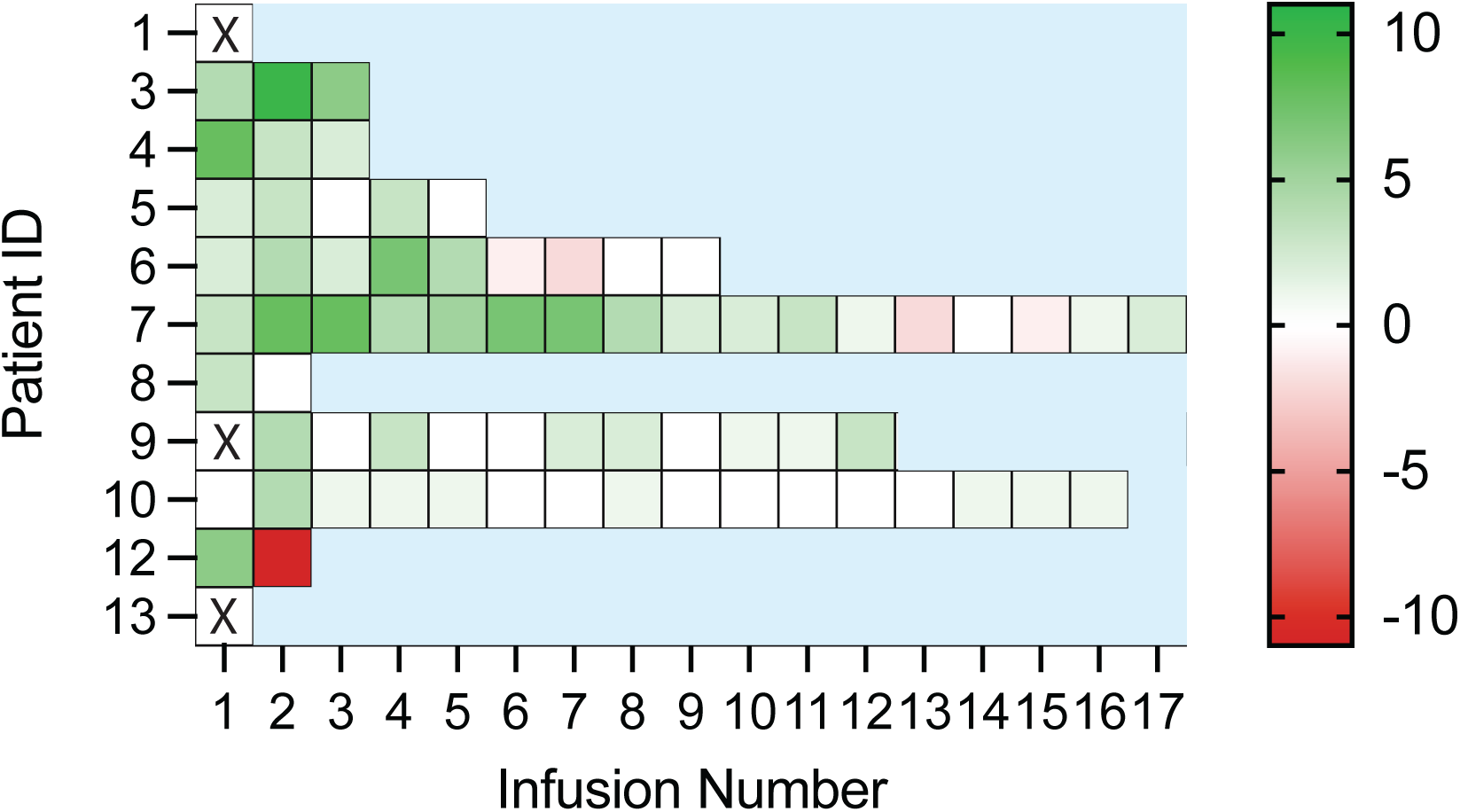
Clinical Improvement Scores. The clinical improvement scale (CIS) is simple tool to quantify changes in the neurological exam^27^ in which each item tested in a comprehensive neurological exam is assigned one point if improved, and -1 point if worse than pre-infusion baseline for each infusion. Clinical improvement scores are assessed one month after each infusion, and compared to the pre-infusion baseline for that infusion. Each GD2-CAR T-cell infusion is treated independently, such that the changes reported compared to the pre-infusion baseline for that infusion. A score of zero (represented as white here) means that there was no clinical change, or that there was an equal number of improved and worsened symptoms/signs following that infusion. Please note that, because the score reflects change after a given infusion, neurological improvement after a previous cycle, with continued benefit but no further improvement, would be annotated with white (no change) in this heatmap. Similarly, improvement in one domain with decreased function in another domain, would also be annotated with white (no change) in this heatmap. Green = better than pre-infusion baseline for that infusion, red = worse than pre-infusion baseline for that infusion.

## Supplemental Tables

**Supplemental Table 1.**
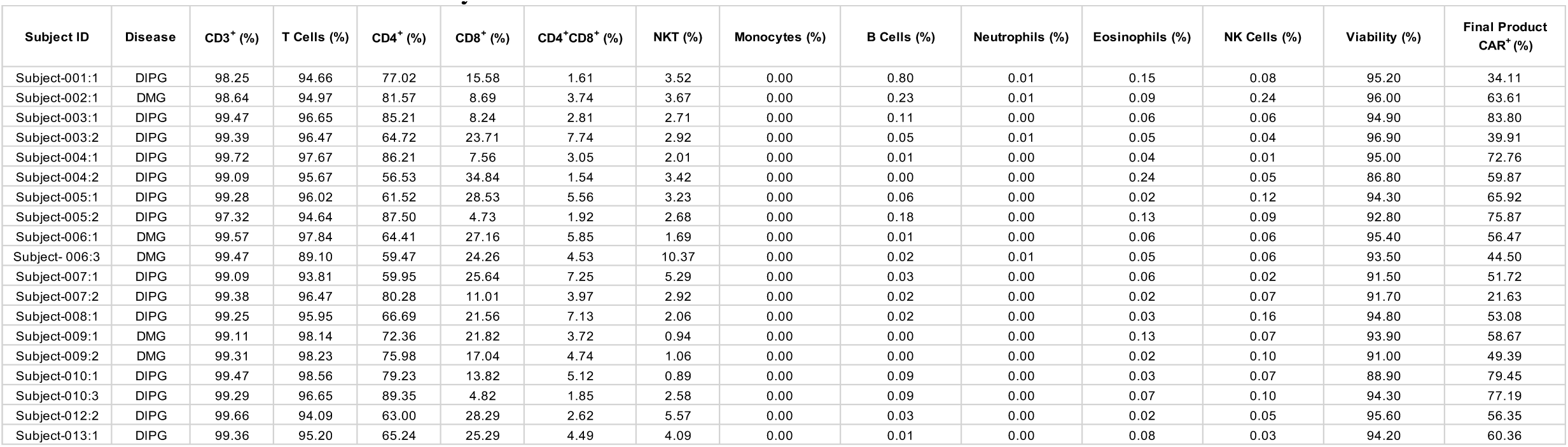
GD2-CAR T drug product phenotypic composition, viability, and transduction efficiency.

**Supplemental Table 2:** CRS, ICANS and TIAN Grading and Management Following IV Infusions.

| Subject | Dose | Max CRS grade | Max ICANS grade | Max TIAN grade | TIAN description* | TIAN management |
| --- | --- | --- | --- | --- | --- | --- |
| 001 | DL1 | 1 | 2 | 4 | Increased ICP to 22 cm H20 (somnolence, worsening of baseline CN VI palsies, dysarthria, ataxia and new CN VII palsy, diplopia, hemiparesis, extensor posturing), impending herniation | dexamethasone, continuous CSF removal through butterfly needle in Ommaya intermittently for four days, hypertonic saline (3%), anakinra |
| 003 | DL1 | 2 | 0 | 2 | Transient worsening of baseline deficits, including trismus and sensory and hearing loss | anakinra |
| 004 | DL1 | 1 | 0 | 2 | Transient worsening of baseline ataxia | anakinra |
| 005 | DL2 | 4 | 0 | 3 | Temporary right diaphragmatic paralysis and worsening of baseline orthostatic hypotension | CPAP, vasopressor support, dexamethasone, anakinra |
| 006^ | DL2 | 3 | 1 | 1 | Severe back pain, worsening paresthesias | anakinra, dexamethasone, pain medications, |
| 007 | DL2 | 2 | 1 | 3 | Increased ICP to 24 cm H20 (somnolence, truncal weakness, worsening of baseline right upper extremity weakness, ataxia and dysarthria) | Continuous CSF removal through butterfly needle in Ommaya for two days, dexamethasone, anakinra |
| 008 | DL2 | 1 | 1 | 3 | Nausea, vomiting, vertigo, altered mental status, increased CN VI and CN VII palsy and R-sided weakness from baseline, small intratumoral/intraventricular hemorrhage | dexamethasone, anakinra |
| 009^ | DL2 | 4 | 3 | 0 |  |  |
| 010 | DL2 | 2 | 0 | 2 | Increased ICP to 19 cm H20, blurry vision, arm paresthesias, ataxic gait | CSF removal with therapeutic tap, anakinra |
| 012 | DL2 | 1 | 0 | 2 | Increased CN III, CN VI, and CN VII palsies from baseline | anakinra |
| 013 | DL2 | 4 | 0 | 4 | Increased ICP to 33 cm H20, increased dysarthria, ataxia, CN VII palsy from baseline | Initially continuous CSF removal through butterfly needle for two days, then endoscopic third ventriculostomy, dexamethasone, anakinra |

Abbreviations: ICP (intracranial pressure), CSF (cerebrospinal fluid), CPAP (continuous airway pressure), CN (cranial nerve)
^ spinal cord diffuse midline glioma
\*All patients experienced headache as part of TIAN

**Supplemental Table 3.**
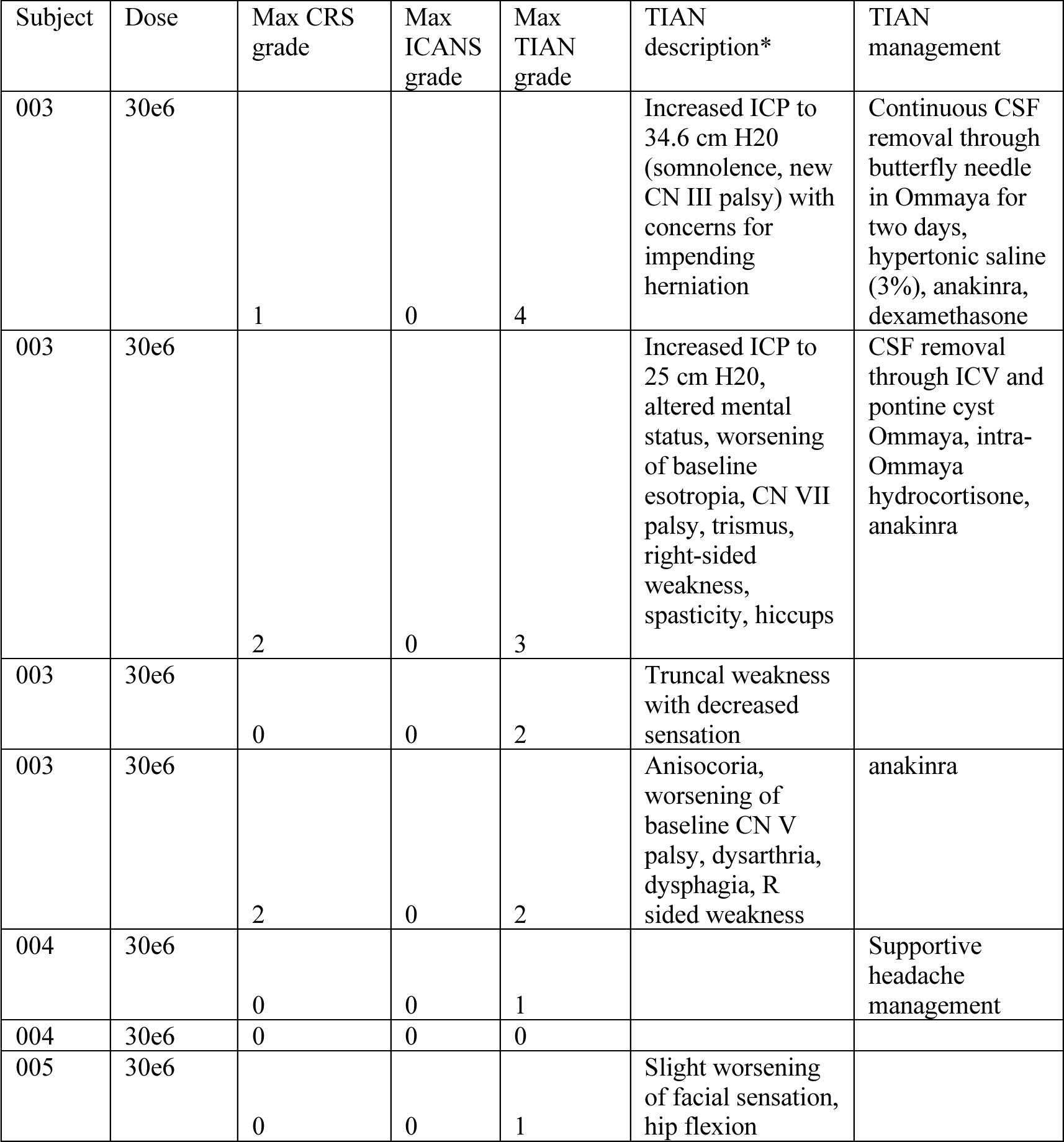
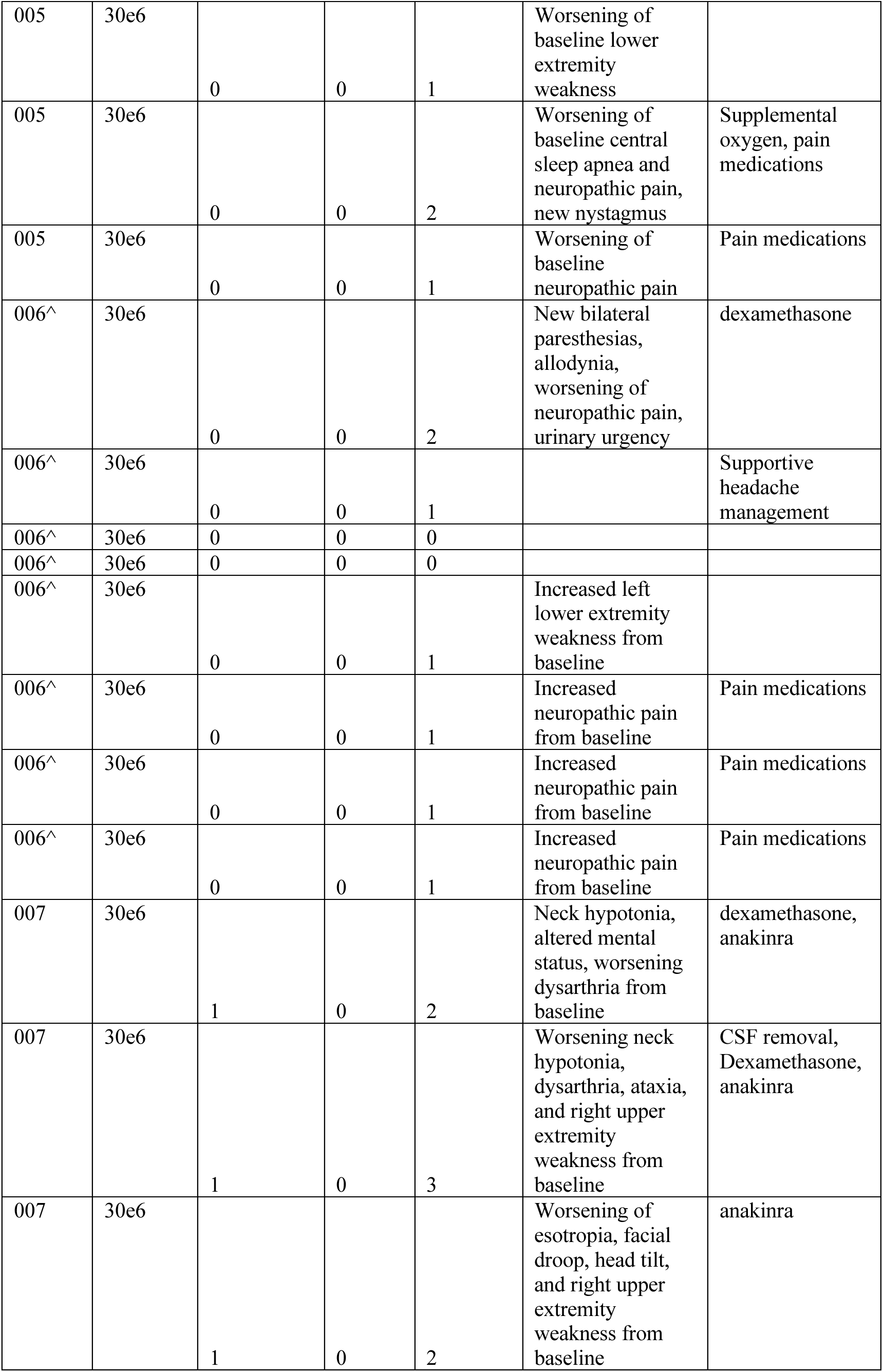
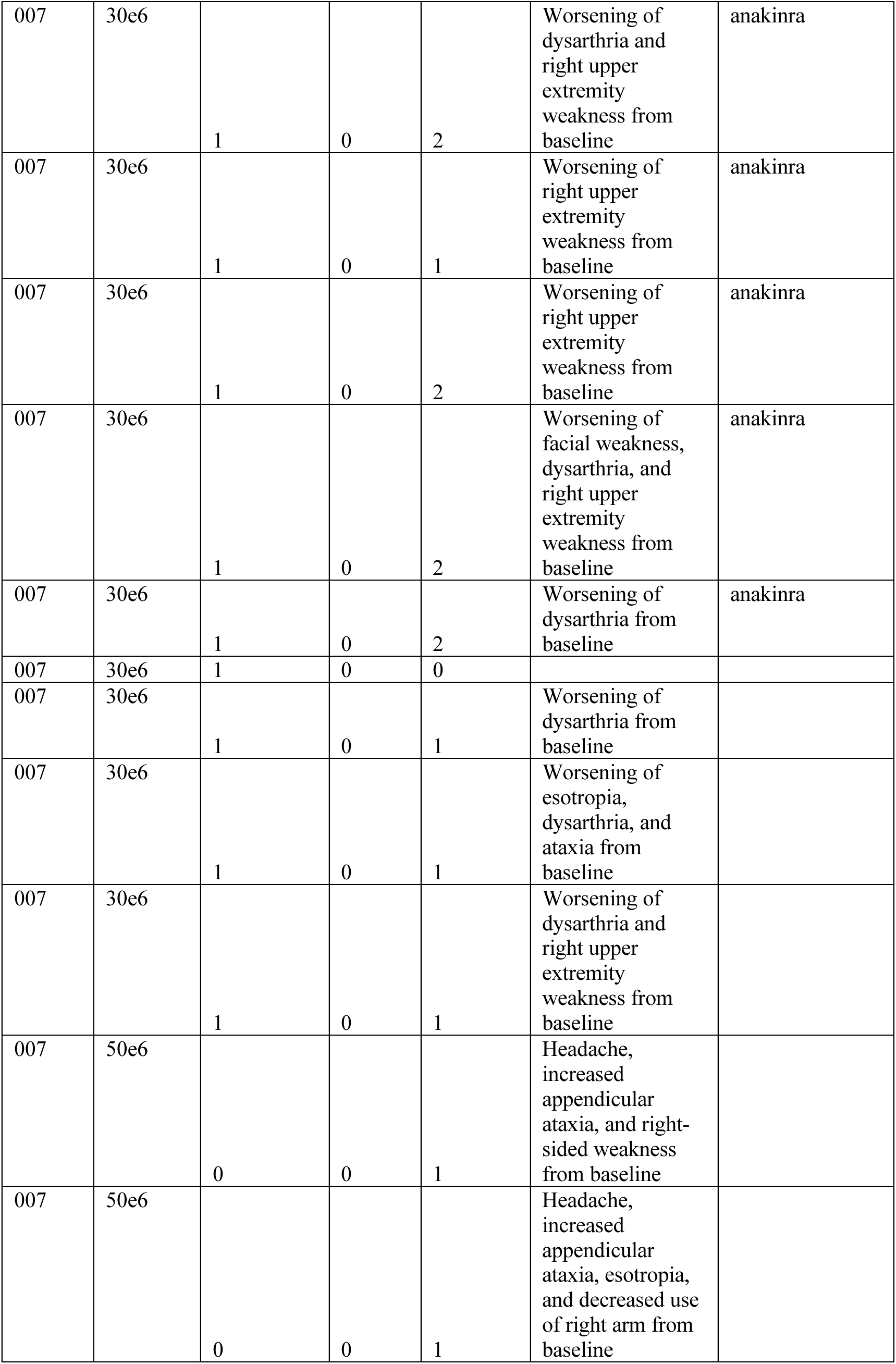
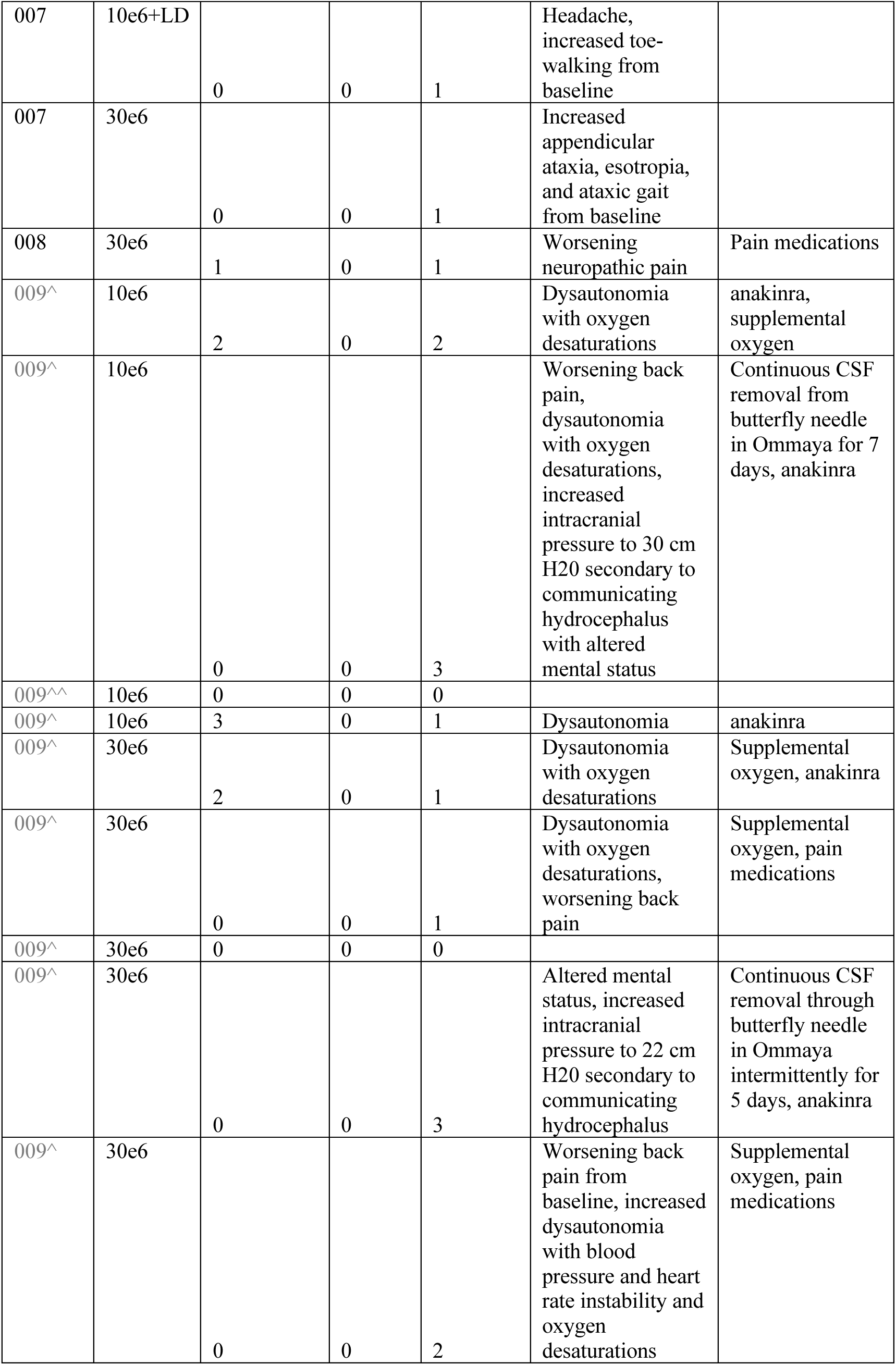
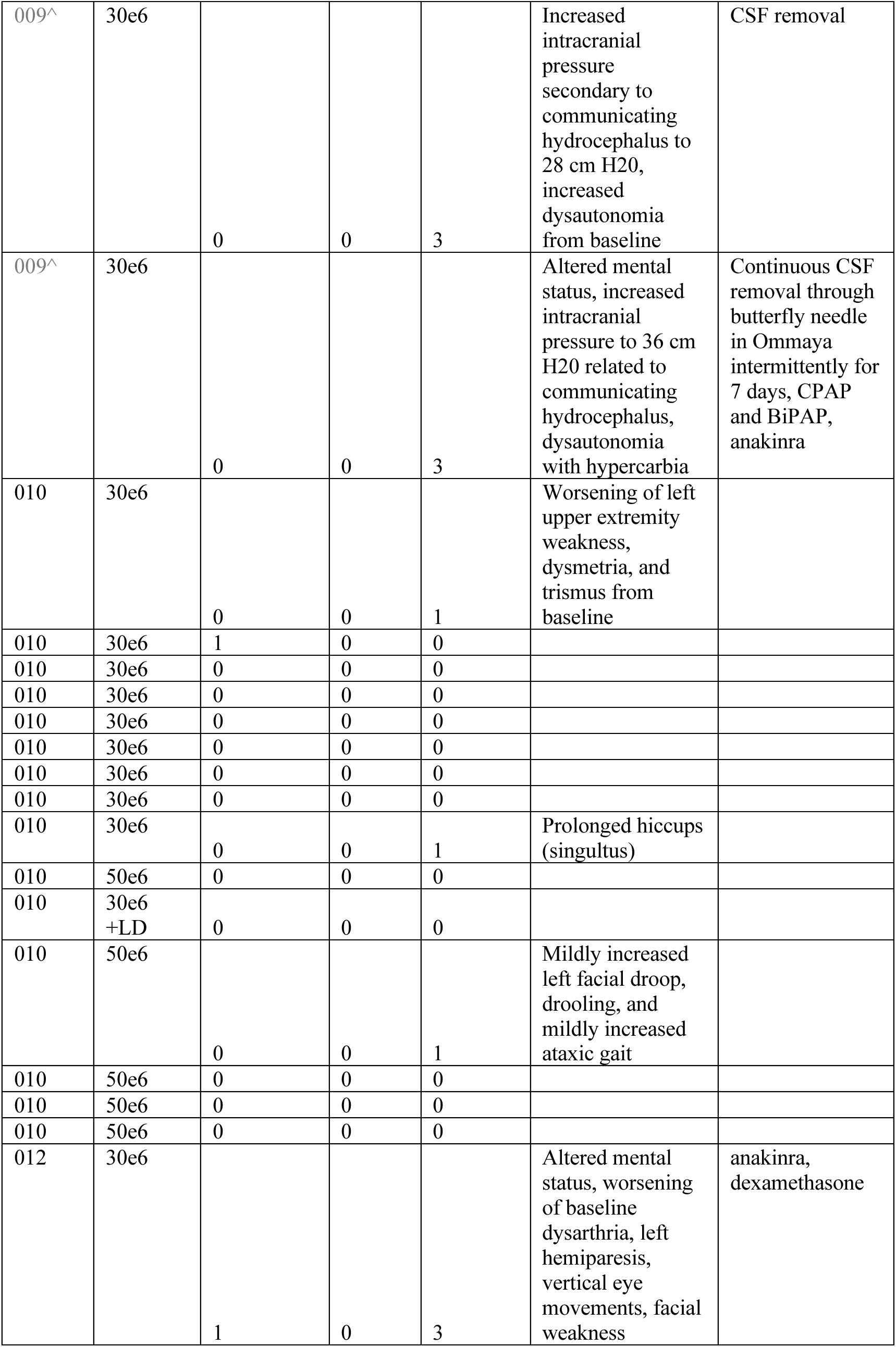
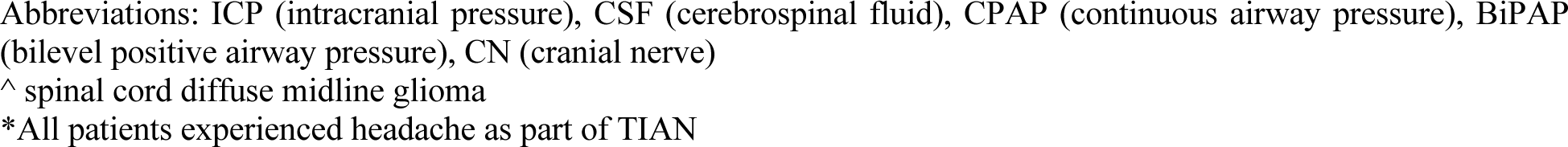
CRS, ICANS and TIAN Grading and Management Following ICV infusions.

**Supplemental Table 4:** Clinical improvement scores.

| Patient | 1st Infusion | 2nd Infusion | 3rd Infusion | 4th Infusion | 5th Infusion | 6th Infusion | 7th Infusion | 8th Infusion | 9th Infusion | 10th Infusion | 11th Infusion | 12th Infusion | 13th Infusion | 14<sup>th</sup> infusion | 15<sup>th</sup> infusion | 16<sup>th</sup> infusion | 17<sup>th</sup> infusion |
| --- | --- | --- | --- | --- | --- | --- | --- | --- | --- | --- | --- | --- | --- | --- | --- | --- | --- |
| 1* |  |  |  |  |  |  |  |  |  |  |  |  |  |  |  |  |  |
| 3 | 4 | 10 | 6 | 1 | 0 |  |  |  |  |  |  |  |  |  |  |  |  |
| 4 | 8 | 3 | 2 |  |  |  |  |  |  |  |  |  |  |  |  |  |  |
| 5 | 2 | 3 | 0 | 3 | 0 |  |  |  |  |  |  |  |  |  |  |  |  |
| 6 | 2 | 4 | 2 | 7 | 4 | -1 | -2 | 0 | 0 |  |  |  |  |  |  |  |  |
| 7 | 3 | 8 | 8 | 4 | 5 | 7 | 7 | 4 | 2 | 2 | 3 | 1 | -2 | 0 | -1 | +1 | +2 |
| 8 | 3 | 0 |  |  |  |  |  |  |  |  |  |  |  |  |  |  |  |
| 9 | * | 4 | 0 | 3 | 0 | 0 | 2 | 2 | 0 | 1 | 1 | 3 |  |  |  |  |  |
| 10 | 0 | 4 | 1 | 1 | 1 | 0 | 0 | 1 | 0 | 0 | 0 | 0 | 0 | +1 | +1 | +1 |  |
| 12 | 6 | -11 |  |  |  |  |  |  |  |  |  |  |  |  |  |  |  |
| 13* |  |  |  |  |  |  |  |  |  |  |  |  |  |  |  |  |  |

\*Not able to assess clinical improvement score because patient was on steroids at the time of clinical evaluation

**Supplemental Table 5:** Grade 3 and 4 Adverse Events.

| Event | Grade 3 Events | Grade 4 Events | Total Events | Patients with Event |
| --- | --- | --- | --- | --- |
| **Cardiovascular disorder** |  |  |  |  |
| Hypertension | 3 |  | 3 | 3 |
| Hypotension | 4 |  | 4 | 4 |
| Sinus tachycardia | 1 |  | 1 | 1 |
| Cytokine release syndrome | 1 | 3 | 4 | 4 |
| **Gastrointestinal disorder** |  |  |  |  |
| Abdominal Pain | 3 |  | 3 | 3 |
| Alanine aminotransferase increased | 3 |  | 3 | 3 |
| Anorexia | 3 |  | 3 | 3 |
| Aspartate aminotransferase increased | 3 |  | 3 | 3 |
| Aspiration |  | 1 | 1 | 1 |
| Constipation | 1 |  | 1 | 1 |
| Dehydration | 1 |  | 1 | 1 |
| Diarrhea | 1 |  | 1 | 1 |
| Gastritis | 1 |  | 1 | 1 |
| Gamma-glutamyl transferase increased | 1 |  | 1 | 1 |
| Nausea | 1 |  | 1 | 1 |
| Lipase increased |  | 1 | 1 | 1 |
| **General disorder** |  |  |  |  |
| Fatigue | 1 |  | 1 | 1 |
| Fever | 6 |  | 6 | 6 |
| Weight loss | 2 |  | 2 | 2 |
| **Hematologic disorder** |  |  |  |  |
| Anemia | 5 |  | 5 | 5 |
| Lymphocyte count decreased | 20 | 18 | 38 | 11 |
| Neutrophil count decreased | 16 | 12 | 28 | 11 |
| Platelet count decreased | 1 |  |  | 1 |
| White blood cell count decreased | 11 | 10 | 21 | 11 |
| Thromboembolic event | 1 |  | 1 | 1 |
| **Infectious disorder** |  |  |  |  |
| Enterocolitis | 1 |  | 1 | 1 |
| Gallbladder infection | 1 |  | 1 | 1 |
| Lung infection | 1 |  | 1 | 1 |
| Sepsis |  | 1 | 1 | 1 |
| Skin infection | 1 |  |  | 1 |
| Urinary tract infection | 5 |  | 5 | 2 |
| Viremia | 1 |  | 1 | 1 |
| **Metabolic disorder** |  |  |  |  |
| Acidosis |  | 1 | 1 | 1 |
| Hypertriglyceridemia |  |  |  |  |
| Hypokalemia | 1 |  | 1 | 1 |
| Hypophosphatemia | 2 |  | 2 | 2 |
| Tumor lysis syndrome | 1 |  | 1 | 1 |
| **Musculoskeletal disorder** |  |  |  |  |
| Back pain | 1 |  | 1 | 1 |
| **Muscle cramp** | 1 |  | 1 | 1 |
| **Muscle weakness** | 4 |  | 4 | 4 |
| **Nervous system disorder** |  |  |  |  |
| **Abducens nerve disorder** | 1 |  | 1 | 1 |
| **Ataxia** | 3 |  | 3 | 3 |
| **Confusion** | 1 |  | 1 | 1 |
| **Dysarthria** | 1 |  | 1 | 1 |
| **Encephalopathy** |  |  |  |  |
| **Facial nerve disorder** | 1 |  | 1 | 1 |
| **Gait disturbance** | 4 |  | 4 | 4 |
| **Glossopharyngeal nerve disorder** | 1 |  | 1 | 1 |
| **Headache** | 3 |  | 3 | 3 |
| **Hydrocephalus** | 8 | 3 | 11 | 6 |
| **Immune effector cell-associated neurotoxicity** | 1 |  | 1 | 1 |
| **Intratumoral hemorrhage** |  |  |  |  |
| **Neck pain** |  |  |  |  |
| **Neuropathic pain** | 1 |  | 1 | 1 |
| **Paresthesia** | 3 |  | 3 | 3 |
| **Phrenic nerve dysfunction** | 1 |  | 1 | 1 |
| **Syncope** | 2 |  | 2 | 2 |
| **Trismus** | 1 |  | 1 | 1 |
| **Tumor inflammation-associated neurotoxicity** | 10 | 3 | 13 | 8 |
| **Urinary incontinence** | 1 |  | 1 | 1 |
| **Vagus nerve dysfunction** | 1 |  | 1 | 1 |
| **Respiratory disorder** |  |  |  |  |
| **Dyspnea** | 1 | 1 | 2 | 2 |
| **Hypoxia** | 2 | 1 | 3 | 3 |
| **Stridor** | 1 |  | 1 | 1 |

## Notes

### Clinical Trial

NCT04196413

### Author Declarations

The IRB of Stanford University gave ethical approval for this work.

## References

1 Maude, S. L. et al. Tisagenlecleucel in Children and Young Adults with B-Cell Lymphoblastic Leukemia. N Engl J Med 378, 439–448 (2018). 10.1056/NEJMoa1709866

2 Schultz, L. M. et al. Disease Burden Affects Outcomes in Pediatric and Young Adult B-Cell Lymphoblastic Leukemia After Commercial Tisagenlecleucel: A Pediatric Real-World Chimeric Antigen Receptor Consortium Report. J Clin Oncol 40, 945–955 (2022). 10.1200/JCO.20.03585

3 Neelapu, S. S. et al. Axicabtagene Ciloleucel CAR T-Cell Therapy in Refractory Large B-Cell Lymphoma. N Engl J Med 377, 2531–2544 (2017). 10.1056/NEJMoa1707447

4 Nastoupil, L. J. et al. Standard-of-Care Axicabtagene Ciloleucel for Relapsed or Refractory Large B-Cell Lymphoma: Results From the US Lymphoma CAR T Consortium. J Clin Oncol 38, 3119–3128 (2020). 10.1200/JCO.19.02104

5 Berdeja, J. G. et al. Ciltacabtagene autoleucel, a B-cell maturation antigen-directed chimeric antigen receptor T-cell therapy in patients with relapsed or refractory multiple myeloma (CARTITUDE-1): a phase 1b/2 open-label study. Lancet 398, 314–324 (2021). 10.1016/S0140-6736(21)00933-8

6 Adusumilli, P. S. et al. A Phase I Trial of Regional Mesothelin-Targeted CAR T-cell Therapy in Patients with Malignant Pleural Disease, in Combination with the Anti-PD-1 Agent Pembrolizumab. Cancer Discov 11, 2748–2763 (2021). 10.1158/2159-8290.CD-21-0407

7 Ahmed, N. et al. HER2-Specific Chimeric Antigen Receptor-Modified Virus-Specific T Cells for Progressive Glioblastoma: A Phase 1 Dose-Escalation Trial. JAMA Oncol 3, 1094–1101 (2017). 10.1001/jamaoncol.2017.0184

8 Ahmed, N. et al. Human Epidermal Growth Factor Receptor 2 (HER2) -Specific Chimeric Antigen Receptor-Modified T Cells for the Immunotherapy of HER2-Positive Sarcoma. J Clin Oncol 33, 1688–1696 (2015). 10.1200/JCO.2014.58.0225

9 Narayan, V. et al. PSMA-targeting TGFbeta-insensitive armored CAR T cells in metastatic castration-resistant prostate cancer: a phase 1 trial. Nat Med 28, 724–734 (2022). 10.1038/s41591-022-01726-1

10 Qi, C. et al. Claudin18.2-specific CAR T cells in gastrointestinal cancers: phase 1 trial interim results. Nat Med 28, 1189–1198 (2022). 10.1038/s41591-022-01800-8

11 Brown, C. E. et al. Locoregional delivery of IL-13Ralpha2-targeting CAR-T cells in recurrent high-grade glioma: a phase 1 trial. Nat Med 30, 1001–1012 (2024). 10.1038/s41591-024-02875-1

12 Choi, B. D. et al. Intraventricular CARv3-TEAM-E T Cells in Recurrent Glioblastoma. N Engl J Med 390, 1290–1298 (2024). 10.1056/NEJMoa2314390

13 Bagley, S. J. et al. Repeated peripheral infusions of anti-EGFRvIII CAR T cells in combination with pembrolizumab show no efficacy in glioblastoma: a phase 1 trial. Nat Cancer 5, 517–531 (2024). 10.1038/s43018-023-00709-6

14 Del Bufalo, F. et al. GD2-CART01 for Relapsed or Refractory High-Risk Neuroblastoma. N Engl J Med 388, 1284–1295 (2023). 10.1056/NEJMoa2210859

15 Cooney, T. et al. Contemporary survival endpoints: an International Diffuse Intrinsic Pontine Glioma Registry study. Neuro Oncol 19, 1279–1280 (2017). 10.1093/neuonc/nox107

16 Mackay, A. et al. Integrated Molecular Meta-Analysis of 1,000 Pediatric High-Grade and Diffuse Intrinsic Pontine Glioma. Cancer Cell 32, 520–537 e525 (2017). 10.1016/j.ccell.2017.08.017

17 Baxter, P. A. et al. A phase I/II study of veliparib (ABT-888) with radiation and temozolomide in newly diagnosed diffuse pontine glioma: a Pediatric Brain Tumor Consortium study. Neuro Oncol 22, 875–885 (2020). 10.1093/neuonc/noaa016

18 Robison, N. J. & Kieran, M. W. Diffuse intrinsic pontine glioma: a reassessment. J Neurooncol 119, 7–15 (2014). 10.1007/s11060-014-1448-8

19 Venneti, S. et al. Clinical efficacy of ONC201 in H3K27M-mutant diffuse midline gliomas is driven by disruption of integrated metabolic and epigenetic pathways. Cancer Discov (2023). 10.1158/2159-8290.CD-23-0131

20 Gallego Perez-Larraya, J., et al. Oncolytic DNX-2401 Virus for Pediatric Diffuse Intrinsic Pontine Glioma. N Engl J Med 386, 2471–2481 (2022). 10.1056/NEJMoa2202028

21 Grassl, N. et al. A H3K27M-targeted vaccine in adults with diffuse midline glioma. Nat Med 29, 2586–2592 (2023). 10.1038/s41591-023-02555-6

22 Mount, C. W. et al. Potent antitumor efficacy of anti-GD2 CAR T cells in H3-K27M(+) diffuse midline gliomas. Nat Med (2018). 10.1038/s41591-018-0006-x

23 Theruvath, J. et al. Locoregionally administered B7-H3-targeted CAR T cells for treatment of atypical teratoid/rhabdoid tumors. Nat Med 26, 712–719 (2020). 10.1038/s41591-020-0821-8

24 Wang, X. et al. The Cerebroventricular Environment Modifies CAR T Cells for Potent Activity against Both Central Nervous System and Systemic Lymphoma. Cancer Immunol Res 9, 75–88 (2021). 10.1158/2326-6066.CIR-20-0236

25 Priceman, S. J. et al. Regional Delivery of Chimeric Antigen Receptor-Engineered T Cells Effectively Targets HER2(+) Breast Cancer Metastasis to the Brain. Clin Cancer Res 24, 95–105 (2018). 10.1158/1078-0432.CCR-17-2041

26 Donovan, L. K. et al. Locoregional delivery of CAR T cells to the cerebrospinal fluid for treatment of metastatic medulloblastoma and ependymoma. Nat Med 26, 720–731 (2020). 10.1038/s41591-020-0827-2

27 Majzner, R. G. et al. GD2-CAR T cell therapy for H3K27M-mutated diffuse midline gliomas. Nature (2022). 10.1038/s41586-022-04489-4

28 Mahdi, J. et al. Tumor inflammation-associated neurotoxicity. Nat Med 29, 803–810 (2023). 10.1038/s41591-023-02276-w

29 Wen, P. Y. et al. RANO 2.0: Update to the Response Assessment in Neuro-Oncology Criteria for High- and Low-Grade Gliomas in Adults. J Clin Oncol, JCO2301059 (2023). 10.1200/JCO.23.01059

30 Kaplan, E. M. P. Nonparametric estimation from incomplete observations. J. Amer. Statist. Assn. 53, 457–481 (1958).

31 Crowley, R. B. J. A Confidence Interval for the Median Survival Time. Biometrics 38, 29–41 (1982).

32 Maude, S. L. et al. Chimeric antigen receptor T cells for sustained remissions in leukemia. N Engl J Med 371, 1507–1517 (2014). 10.1056/NEJMoa1407222

33 Lee, D. W. et al. T cells expressing CD19 chimeric antigen receptors for acute lymphoblastic leukaemia in children and young adults: a phase 1 dose-escalation trial. Lancet 385, 517–528 (2015). 10.1016/S0140-6736(14)61403-3

34 Fry, T. J. et al. CD22-targeted CAR T cells induce remission in B-ALL that is naive or resistant to CD19-targeted CAR immunotherapy. Nat Med 24, 20–28 (2018). 10.1038/nm.4441

35 Mueller, S. et al. PNOC015: Repeated convection enhanced delivery (CED) of MTX110 (aqueous panobinostat) in children with newly diagnosed diffuse intrinsic pontine glioma (DIPG). Neuro Oncol (2023). 10.1093/neuonc/noad105

36 Mueller, S. et al. Mass cytometry detects H3.3K27M-specific vaccine responses in diffuse midline glioma. J Clin Invest 130, 6325–6337 (2020). 10.1172/JCI140378

37 Gardner, S. L. et al. Phase I dose escalation and expansion trial of single agent ONC201 in pediatric diffuse midline gliomas following radiotherapy. Neurooncol Adv 4, vdac143 (2022). 10.1093/noajnl/vdac143

38 Roberts, H. J. et al. Clinical, genomic, and epigenomic analyses of H3K27M-mutant diffuse midline glioma long-term survivors reveal a distinct group of tumors with MAPK pathway alterations. Acta Neuropathol 146, 849–852 (2023). 10.1007/s00401-023-02640-7

39 Pathania, M. et al. H3.3(K27M) Cooperates with Trp53 Loss and PDGFRA Gain in Mouse Embryonic Neural Progenitor Cells to Induce Invasive High-Grade Gliomas. Cancer Cell 32, 684–700 e689 (2017). 10.1016/j.ccell.2017.09.014

